# Time trends in social contacts of individuals according to comorbidity and vaccination status, before and during the COVID-19 pandemic

**DOI:** 10.1101/2021.12.02.21267205

**Authors:** Aurélie Godbout, Mélanie Drolet, Myrto Mondor, Marc Simard, Chantal Sauvageau, Gaston De Serres, Marc Brisson

## Abstract

**Background:** As we are confronted with more transmissible/severe variants with immune escape and the waning of vaccine efficacy, it is particularly relevant to understand how the social contacts of individuals at greater risk of COVID-19 complications evolved over time. We described time trends in social contacts of individuals according to comorbidity and vaccination status before and during the first three waves of the COVID-19 pandemic in Quebec, Canada.

**Methods:** We used data from CONNECT, a repeated cross-sectional population-based survey of social contacts conducted before (2018/2019) and during the pandemic (April 2020 to July 2021). We recruited non-institutionalized adults from Quebec, Canada, by random digit dialling. We used a self-administered web-based questionnaire to measure the number of social contacts of participants (two-way conversation at a distance ≤2 meters or a physical contact, irrespective of masking). We compared the mean number of contacts/day according to the comorbidity status of participants (pre-existing medical conditions with symptoms/medication in the past 12 months) and 1-dose vaccination status during the third wave. All analyses were performed using weighted generalized linear models with a Poisson distribution and robust variance.

**Results:** A total of 1441 and 5185 participants with and without comorbidities, respectively, were included in the analyses. Contacts significantly decreased from a mean of 6.1 (95%CI 4.9–7.3) before the pandemic to 3.2 (95%CI 2.5–3.9) during the first wave among individuals with comorbidities, and from 8.1 (95%CI 7.3–9.0) to 2.7 (95%CI 2.2–3.2) among individuals without comorbidities. Individuals with comorbidities maintained fewer contacts than those without comorbidities in the second wave, with a significant difference before the Christmas 2020/2021 holidays (2.9 (95%CI 2.5–3.2) *v* 3.9 (95%CI 3.5–4.3); P<0.001). During the third wave, contacts were similar for individuals with (4.1, 95%CI 3.4– 4.7) and without comorbidities (4.5, 95%CI 4.1–4.9; P=0.27). This could be partly explained by individuals with comorbidities vaccinated with their first dose who increased their contacts to the level of those without comorbidities.

**Conclusions:** It will be important to closely monitor COVID-19-related outcomes and social contacts by comorbidity and vaccination status to inform targeted or population-based interventions (e.g., booster doses of the vaccine).

## BACKGROUND

Since the beginning of the COVID-19 pandemic, most countries have introduced unprecedented physical distancing measures to slow down transmission. Several studies have shown that these measures were associated with substantial decreases of social contacts in the general population [1-3], which contributed to flattening the epidemic curves while awaiting safe and efficacious vaccines [4, 5]. We also learned from data collected during the first wave that individuals with comorbidities (e.g., obesity, diabetes, heart disease) were at greater risk of complications from COVID-19. Age-adjusted relative risks of hospitalisation and death were up to 1.5 times higher among individuals with comorbidities compared to those without comorbidities [6]. However, we do not know whether there was a differential evolution of social contacts according to the risk of COVID-19 complications. If so, individuals who felt at greater risk of complications could have shielded themselves more by having fewer contacts, thereby reducing the severity of COVID-19 in terms of hospitalisation and death per case at the population-level. Recent data from Quebec, Canada, showed that there were proportionately fewer cases of COVID-19 with comorbidities in the second wave (43%) compared to the first wave (49%) when excluding nursing home cases (Additional file 1: Table S1) [6]. This might have contributed to the decrease in the average age-adjusted severity of COVID-19 observed during the second wave compared to the first wave [7-9].

In December 2020, one year after the beginning of the pandemic, the first COVID-19 vaccine was shown to be safe and highly efficacious against the disease and its complications [10]. Given limited vaccine supply, most countries initially started with targeted vaccination of groups at greater risk of complications (e.g., individuals living in long-term care facilities and individuals with comorbidities) and greater risk of exposure/transmission (e.g., health care workers, essential workers) before expanding vaccination to the general population, starting with the oldest age groups [11-14]. As vaccination coverage increased, physical distancing measures could be gradually relaxed, allowing for increased social contacts [15-17]. To our knowledge, no study has documented how individuals’ vaccination status influences their contacts, particularly for those at greater risk of COVID-19 complications.

Although high vaccine efficacy against COVID-19 was observed in randomized clinical trials for all age groups and for individuals at risk or not for severe COVID-19 [10, 18], recent population-level data suggest a potential waning of vaccine effectiveness over time and vaccine escape with the Omicron variant [19-24]. For example, studies conducted in United Kingdom and Qatar showed decreased effectiveness of COVID-19 vaccines against infection five months after the second dose [19, 20]. In the UK study, the waning of vaccine effectiveness against symptomatic COVID-19 was more important for individuals aged 65 years or more and for individuals with underlying medical conditions [19]. However, limited waning of two-dose vaccine effectiveness against COVID-19-related hospitalisations and deaths was observed in both studies and in Canada, before the Omicron wave [19, 20, 25]. More recently, a decline in vaccine effectiveness against Omicron-related infections and hospitalisations was observed because of its high vaccine escape [22-24]. In response to this possible decrease in vaccine effectiveness over time and the emergence of Omicron, some countries have introduced booster doses for individuals at greater risk of complications [26-29]. Understanding how the social contacts of individuals at greater risk of complications evolved over time, particularly after vaccination, is therefore highly relevant in this context.

Examining time trends in social contacts according to the risk of complications of individuals can provide insight into the differences in severity of the different waves and help inform public-health decisions about the need for future preventive measures for individuals at greater risk of complications, who may have increased their contacts after vaccination. This information is particularly relevant as we are confronted with more transmissible/severe variants (e.g., Delta) with high immune escape (e.g., Omicron) and with the waning of vaccine efficacy. The main objective of this study is to compare time trends in social contacts of Quebec adults with and without comorbidities before the pandemic and during the first three waves of the pandemic. A secondary objective is to explore whether vaccination with the first dose of the COVID-19 vaccine influenced the association between social contacts and comorbidities.

## METHODS

This paper was written according to the Strengthening the Reporting of Observational Studies in Epidemiology (STROBE) statement (Additional file 1: Table S2) [30].

### Study design

We used data from a repeated cross-sectional population-based survey of social contacts (CONNECT – CONtact and Network Estimation to Control Transmission) conducted before and during the COVID-19 pandemic in Quebec, Canada. The detailed methodology of CONNECT has been described previously [3]. The first phase was conducted from February 2018 to March 2019, one year before the COVID-19 pandemic. In order to document the changes of social contacts during the pandemic, additional phases were undertaken (April 21^st^-May 25^th^ 2020 and July 3^rd^ 2020-July 4^th^ 2021). The same methodology was used for all CONNECT phases.

### Recruitment of participants

All non-institutionalized Quebecers without age limits were targeted for CONNECT recruitment. We restricted the current analyses to participants aged 18 years or older because they represent the vast majority of COVID-19 hospitalisations and deaths [7, 8]. We recruited participants by random digit dialling, using landline and mobile phone numbers. First, we explained the study to the respondent, verified the household eligibility, and documented the age and sex of all household members. Then, we used an age-stratified probability sampling to randomly select one individual per household to participate in CONNECT. We recruited new participants for each phase of CONNECT using the same procedure. The CONNECT study was approved by the ethics committee of the CHU de Québec research center and participants gave their consent to participate in the study during the recruitment phone call. We commissioned the market company Advanis for the recruitment of participants and data collection.

### Data collection

We used a self-administered online questionnaire for data collection and the same questionnaire was used for all CONNECT phases. After the selected participants gave their consent to participate in the study, we sent them an email containing a secured individualized web link to the questionnaire and additional information about the study.

In the first section of the questionnaire, we documented key socio-demographic characteristics (e.g., age, sex, region, household size, education level, country of origin, race/ethnicity, main occupation) and the health condition of participants. Specifically, participants were questioned about any long-term health condition(s), which is expected to last or has already lasted 6 months or more, diagnosed by a health care professional they may have or have had. For each condition reported, they were asked whether they have had symptoms or taken medication in the past 12 months (Additional file 1: Example of questions S1). The second section of the questionnaire was a social contact diary, adapted from Polymod and other similar studies (Additional file 1: Example of questions S2) [31-33]. We assigned each participant two random days of the week (one week day and one weekend day) to document every different person they had contact with between 5 am and 5 am the following morning. We defined contacts as either physical (handshake, hug, kiss) or nonphysical (two-way conversation in the physical presence of the person at a distance equal or less than 2 meters, irrespective of masking). Participants recorded in the diary the characteristics of the contact person (age, sex, race/ethnicity, and relationship to themselves) and the characteristics of the contact itself (location (home, work, school, public transportation, leisure, other locations), duration, frequency, and whether the contact was physical or not). When participants reported having more than 20 professional contacts per working day, we asked them general questions about these professional contacts (age groups of the majority of contact persons, average duration of contacts, and whether physical contacts were generally involved or not) rather than reporting each professional contact in the diary. Since the beginning of January 2021, we asked participants whether they have been vaccinated against COVID-19 and, if so, the date(s) of vaccination.

### Main outcome and exposure variables

Our main outcome was the mean number of social contacts per person and per day, for all locations combined. We weighted contacts reported on weekdays (5/7) and the weekend (2/7) to represent the mean number of social contacts per day over a week. To distinguish between contacts at home with household members from the other contacts that could be influenced by physical distancing measures, we also stratified contacts into two groups: 1) contacts at home with household members, and 2) contacts at home with visitors and contacts in all other locations. If a contact was reported both at home and in another location, we only considered the contact at home, where the risk of transmission is higher, to avoid counting multiple contacts with the same person. As other previous studies that limited the number of contacts per day [31, 32], we truncated professional contacts to a maximum of 40 per day to eliminate contacts at low risk of infectious disease transmission and extreme values.

Our main exposure variable was the presence of an active physical comorbidity, that is a long-term physical health condition for which participants have had symptoms or taken medication in the past 12 months (excluding mental health conditions). We restricted the sensitivity analyses to active physical comorbidities that were shown to increase the risk of COVID-19 complications according to two analyses, one from Institut national de santé publique du Québec (INSPQ) and another one from National Advisory Committee on Immunization (NACI) (Additional file 1: Table S3 and Figure S1) [6, 34]. Active physical comorbidities are referred thereafter in the text as comorbidities.

For the vaccination status, participants were considered vaccinated if they reported receiving their first dose of COVID-19 vaccine before or at their assigned day for the study. Although randomized control trials have shown high vaccine efficacy 14 days after vaccination [10, 18], we did not consider an interval after vaccination because anecdotal reports suggested that individuals may feel protected immediately after vaccination [35]. The two assigned days were considered separately for the vaccination status, meaning that if participants were vaccinated in the interval between their first and second assigned day, they were considered vaccinated only for the second day.

### Analyses

We weighted CONNECT participants by age, sex, region (Greater Montreal and other Quebec regions), and household composition, using the 2016 Canadian census data of Quebec [36]. We performed all analyses using weighted generalized linear models with a Poisson distribution, and an identity link was used to obtain mean differences. To take into account the correlation between the two assigned days of each participant and data overdispersion, we used generalized estimating equations with robust variance [37]. Participants with inadequately completed social contact diaries or with missing values for comorbidities were excluded from the analyses.

For the main analysis, we compared the mean number of social contacts per day of individuals with and without comorbidities, for the pre-pandemic period and the first three waves of COVID-19. Periods were determined according to COVID-19 epidemiology in Quebec and CONNECT data availability: pre-COVID in 2018-2019, first wave from April 21^st^ to May 25^th^ 2020, summer 2020 from July 3^rd^ to August 22^nd^, second wave from August 23^rd^ 2020 to March 20^th^ 2021, and third wave from March 21^st^ to July 4^th^ 2021. The second wave was further stratified to consider the Christmas holidays separately: before the holidays from August 23^rd^ to December 16^th^ 2020, holidays from December 17^th^ 2020 to January 8^th^ 2021 and after the holidays from January 9^th^ to March 20^th^ 2021 (Figure 1). We performed the analyses for all adults and stratified by age (18 to 65 and over 65 years old). We considered the following potentially confounding variables: age, sex, region, household size, education level, main occupation, race/ethnicity, and country of origin. Age was the only confounding variable identified using the change in estimate method with backward selection [38, 39]. Therefore, we adjusted analyses for age (18 to 25, 26 to 45, 46 to 65, over 65 years old). Stratified analyses on the subgroup of participants aged over 65 years were not adjusted since there were too few participants for the adjustment. In sensitivity analysis, we replicated the same analysis, but using the classifications of comorbidities at risk of COVID-19 complications from INSPQ and NACI. Given that comparisons of social contacts between individuals with and without comorbidities represent a secondary analysis of the CONNECT study, we calculated the power using the number of participants available for these analyses and the proportion with comorbidities. For example, with a sample size of 1200 individuals, 22% of which having comorbidities, it is possible to detect a difference of 0.9 contact (3.0 vs 3.9) with 85% power and 5% two-tailed type I error, taking into account a design effect of 5 due to overdispersion and correlation between days (Additional file 1: Table S4).

**Figure 1.**
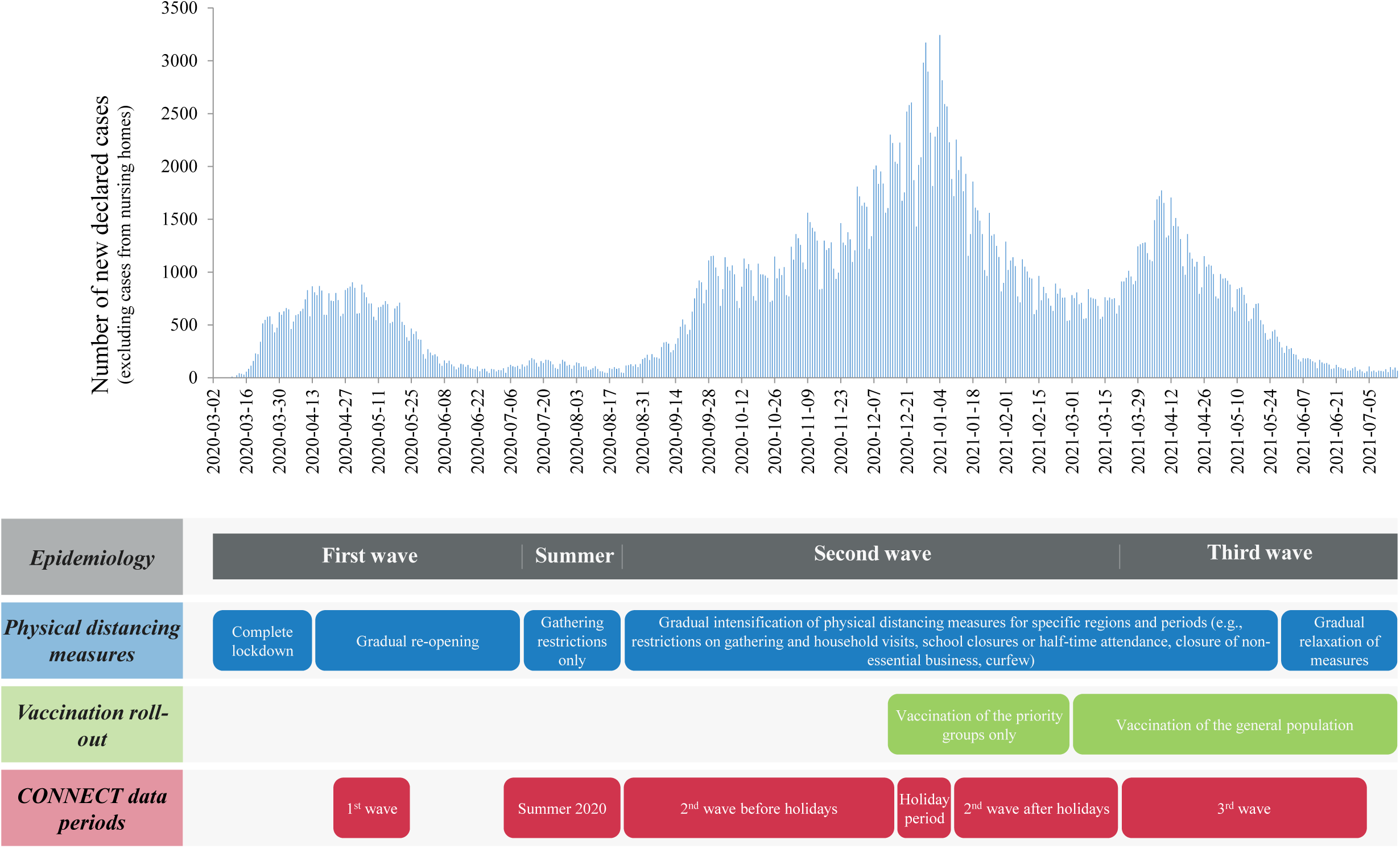
Timeline of COVID-19 epidemiology, physical distancing measures and vaccination roll-out in Quebec in relation to CONNECT data periods [57].

For the secondary analysis, we compared the mean number of social contacts per day of individuals with and without comorbidities during the third wave, according to their vaccination status. This analysis was stratified by age and adjusted for time periods (March, April, May, June-July) since vaccination status and contacts varied substantially during the third wave as a result of the general population vaccination roll-out starting in March 2021 and gradual relaxation of physical distancing measures (Figure 1). All statistical analyses were performed using SAS version 9.4.

## RESULTS

### Participants

A total of 6836 adults from Quebec completed the CONNECT questionnaire and 210 were excluded as a result of inadequately completed social contact diary or missing values related to comorbidity status. The current analysis included 6626 adults (1441 and 5185 with and without comorbidities, respectively) (Table 1). Participants with comorbidities were older (35% over 65 years old) than those without comorbidities (17% over 65 years old), they lived in smaller households (1-2 people) and were more likely to be retired or unemployed. Participants with and without comorbidities were similar for the other socio-demographic characteristics and were comparable across CONNECT periods (Additional file 1: Table S5). The majority of participants with comorbidities had one comorbidity (78%) and the most common comorbidities were chronic lung disease and diabetes (Table 1 and Additional file 1: Table S5).

**Table 1.** Key socio-demographic characteristics of participants.

|  | All participants |  | With comorbidities |  | Without comorbidities |  |
| --- | --- | --- | --- | --- | --- | --- |
|  | N<sub>crude</sub> | % weighted | N<sub>crude</sub> | % weighted | N<sub>crude</sub> | % weighted |
| **Total** | **6626** |  | **1441** |  | **5185** |  |
| **Age** |  |  |  |  |  |  |
| 18-25 yrs old | 865 | 11.7 | 69 | 3.6 | 796 | 14.1 |
| 26-45 yrs old | 2204 | 34.5 | 352 | 23.8 | 1852 | 37.6 |
| 46-65 yrs old | 2503 | 33.0 | 644 | 37.7 | 1859 | 31.7 |
| 66-75 yrs old | 901 | 12.5 | 305 | 18.7 | 596 | 10.6 |
| >75 yrs old | 153 | 8.3 | 71 | 16.3 | 82 | 5.9 |
| **Sex** |  |  |  |  |  |  |
| Male | 3114 | 49.9 | 670 | 48.9 | 2444 | 50.2 |
| Female | 3512 | 50.1 | 771 | 51.1 | 2741 | 49.8 |
| **Region** |  |  |  |  |  |  |
| Greater Montreal<sup>†</sup> | 3805 | 61.6 | 835 | 62.4 | 2970 | 61.3 |
| Other Quebec regions | 2808 | 38.5 | 605 | 37.6 | 2203 | 38.7 |
| Missing | 13 | -- | 1 | -- | 12 | -- |
| **Household size** |  |  |  |  |  |  |
| 1 | 1695 | 26.3 | 433 | 30.9 | 1262 | 25.0 |
| 2 | 3281 | 47.7 | 789 | 53.9 | 2492 | 45.9 |
| 3 | 789 | 11.7 | 107 | 7.2 | 682 | 13.1 |
| 4+ | 861 | 14.3 | 112 | 8.1 | 749 | 16.1 |
| **Education level** |  |  |  |  |  |  |
| No diploma, degree | 301 | 4.6 | 83 | 5.6 | 218 | 4.3 |
| Secondary (high) school | 890 | 13.5 | 192 | 13.2 | 698 | 13.6 |
| College, CEGEP, university or other certificate/diploma | 5435 | 82.0 | 1166 | 81.2 | 4269 | 82.2 |
| **Main occupation** |  |  |  |  |  |  |
| Student employed or unemployed | 761 | 10.8 | 84 | 5.1 | 677 | 12.5 |
| Employed or semi-retired | 3794 | 55.0 | 688 | 43.1 | 3106 | 58.5 |
| Temporarily not working or seeking work | 437 | 5.9 | 118 | 7.2 | 319 | 5.6 |
| Unemployed or retired | 1634 | 28.3 | 551 | 44.6 | 1083 | 23.4 |
| **Type of employment (among 18-65 yrs old employed or temporarily not working)** |  |  |  |  |  |  |
| Education | 399 | 9.4 | 75 | 9.7 | 324 | 9.4 |
| Health | 379 | 8.7 | 79 | 9.7 | 300 | 8.5 |
| Sales and services | 537 | 12.9 | 98 | 13.6 | 439 | 12.8 |
| Other sectors | 2802 | 69.0 | 490 | 67.0 | 2312 | 69.4 |
| Missing | 23 | -- | 3 | -- | 20 | -- |
|  | N<sub>crude</sub> | % weighted | N<sub>crude</sub> | % weighted | N<sub>crude</sub> | % weighted |
| **Race/Ethnicity** |  |  |  |  |  |  |
| White | 5983 | 91.2 | 1337 | 94.0 | 4646 | 90.4 |
| Other | 561 | 8.8 | 88 | 6.0 | 473 | 9.6 |
| Missing | 82 | -- | 16 | -- | 66 | -- |
| **Country of origin** |  |  |  |  |  |  |
| Canadian-born | 5917 | 88.7 | 1330 | 91.8 | 4587 | 87.7 |
| Foreign-born | 700 | 11.3 | 111 | 8.2 | 589 | 12.3 |
| Missing | 9 | -- | -- | -- | 9 | -- |
| **Number of comorbidities** |  |  |  |  |  |  |
| 1 | -- | -- | 1128 | 78.4 | -- | -- |
| 2 | -- | -- | 246 | 16.9 | -- | -- |
| 3+ | -- | -- | 67 | 4.7 | -- | -- |
| **Type of comorbidities** |  |  |  |  |  |  |
| Chronic lung disease | -- | -- | 315 | 20.6 | -- | -- |
| Diabetes | -- | -- | 308 | 21.3 | -- | -- |
| Chronic inflammatory disease | -- | -- | 236 | 16.0 | -- | -- |
| Hypertension | -- | -- | 165 | 12.8 | -- | -- |
| Chronic heart disease | -- | -- | 137 | 10.3 | -- | -- |
| Cancer | -- | -- | 98 | 8.2 | -- | -- |
| Thyroid disease | -- | -- | 96 | 6.9 | -- | -- |
| Neurologic disease | -- | -- | 89 | 5.4 | -- | -- |
| Arthritis and arthrosis | -- | -- | 65 | 4.5 | -- | -- |
| Others | -- | -- | 303 | 19.9 | -- | -- |

† Greater Montreal: Regions of Montreal, Laval, Montérégie, Lanaudière, Laurentides

### Time trends in social contacts of individuals with and without comorbidities

During the pre-pandemic period, individuals with comorbidities had significantly fewer contacts per day (mean 6.1, 95%CI 4.9–7.3) than those without comorbidities (8.1, 95%CI 7.3–9.0; P=0.008) (Figure 2, Additional file 1: Table S6A). The mean number of contacts decreased significantly during the first wave of COVID-19, to 3.2 (95%CI 2.5–3.9) and 2.7 (95%CI 2.2–3.2) for individuals with and without comorbidities, respectively, and there was no significant difference between the two groups (P=0.23). Contacts then increased significantly for both groups during summer 2020 to 4.2 (95%CI 3.5–4.9) and 4.3 (95%CI 3.6–5.0). During the second wave, from the end of August 2020 to March 2021, individuals with comorbidities maintained fewer contacts than individuals without comorbidities, with a significant difference between the two groups before the Christmas holidays (2.9 (95%CI 2.5–3.2) *v* 3.9 (95%CI 3.5–4.3); P<0.001). However, during the holidays, individuals without comorbidities significantly decreased their contacts to 2.8 (95%CI 2.5–3.1) because of school/work vacations and gathering restrictions, and no difference was observed during this period compared to individuals with comorbidities (3.0, 95%CI 2.6–3.4; P=0.41). During the third wave, contacts increased significantly to 4.1 (95%CI 3.4–4.7) and 4.5 (95%CI 4.1–4.9) for individuals with and without comorbidities, respectively, and there was no significant difference between the two groups (P=0.27). When using the other two classifications of comorbidities for the sensitivity analyses, similar results were observed (Additional file 1: Table S7).

**Figure 2.**
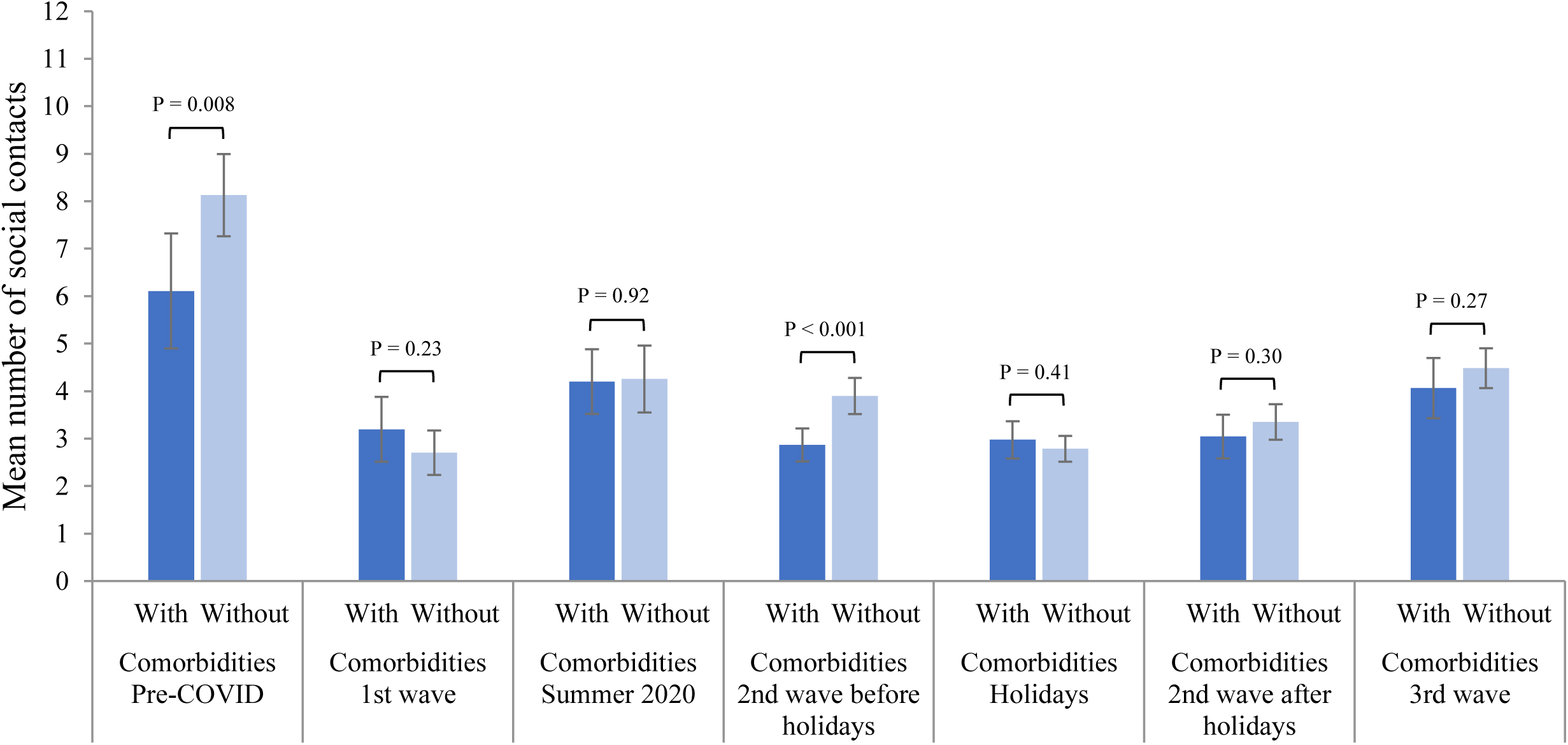
Time trends in the mean total number of social contacts of individuals with and without comorbidities*. **Pre-COVID:** February 1^st^ 2018 to March 17^th^ 2019; **1**^**st**^ **wave:** April 21^st^ to May 25^th^ 2020; **Summer 2020:** July 3^rd^ to August 22^nd^ 2020; **2**^**nd**^ **wave:** August 23^rd^ 2020 to March 20^th^ 2021; **Holidays:** December 17^th^ 2020 to January 8^th^ 2021; **3**^**rd**^ **wave:** March 21^st^ to July 4^th^ 2021. Results are presented with 95% confidence intervals and p-values of the difference between individuals with and without comorbidities. *Results adjusted for age

Time trends were generally the same as previously described among individuals aged 18 to 65 years and those aged over 65 years from the pre-pandemic period throughout the Christmas holidays (Figure 3, Additional file 1: Table S6A). However, during the second wave after the holidays, individuals aged 18 to 65 years with comorbidities maintained significantly fewer contacts (2.8, 95%CI 2.1–3.6) than those without comorbidities (3.9, 95%CI 3.3–4.4; P=0.03). In contrast, there was no significant difference between individuals over 65 years old with (1.6, 95%CI 1.1–2.1) and without comorbidities (1.5, 95%CI 1.1–2.0; P=0.87) during that period. No significant difference was observed between individuals with and without comorbidities aged 18 to 65 years and over 65 years during the third wave.

**Figure 3.**
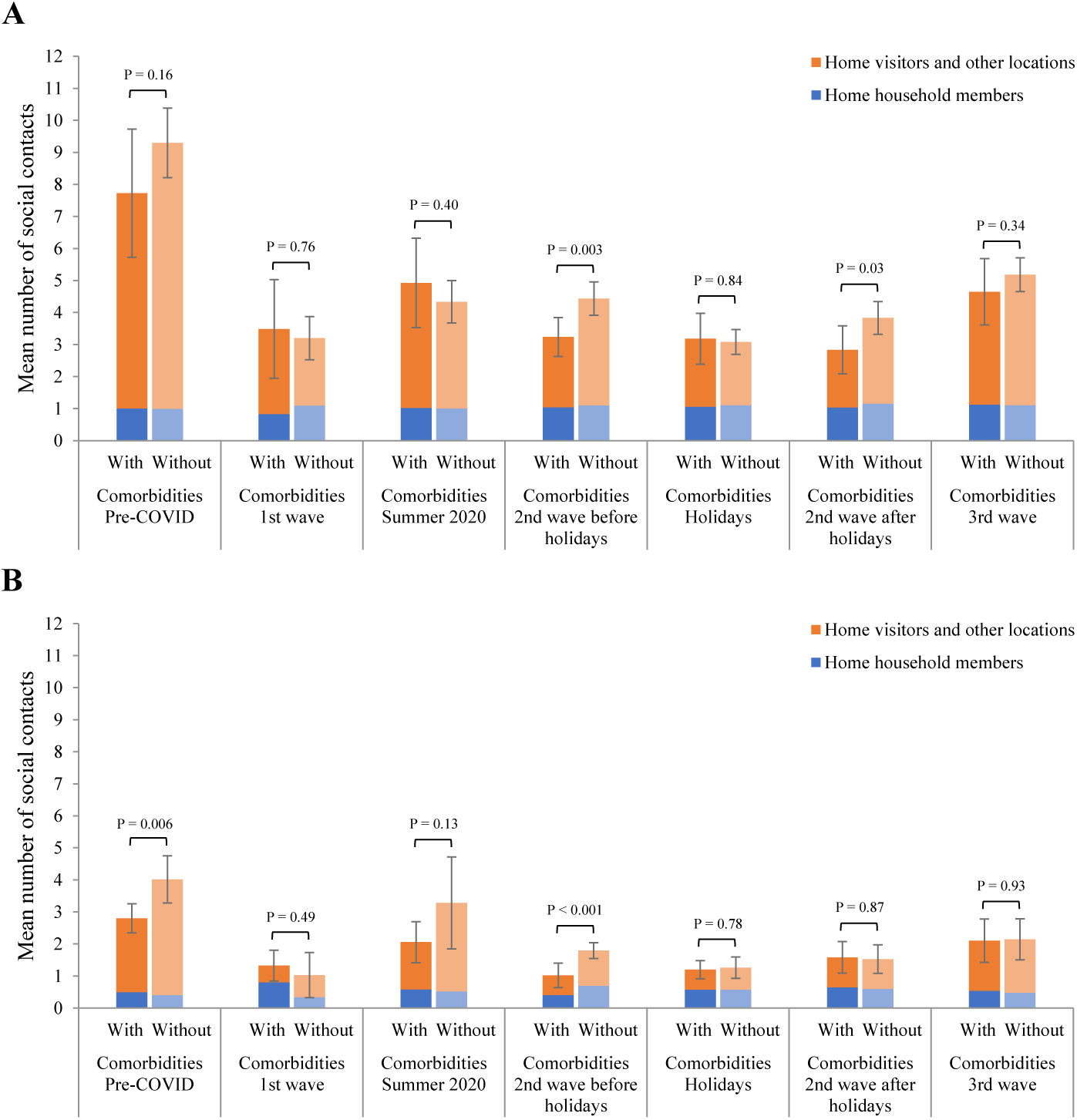
Time trends in the mean number of social contacts of individuals with and without comorbidities, by age and contact location. **A) Adults 18 to 65 years old\*** **Pre-COVID:** February 1^st^ 2018 to March 17^th^ 2019; **1**^**st**^ **wave:** April 21^st^ to May 25^th^ 2020; **Summer 2020:** July 3^rd^ to August 22^nd^ 2020; **2**^**nd**^ **wave:** August 23^rd^ 2020 to March 20^th^ 2021; **Holidays:** December 17^th^ 2020 to January 8^th^ 2021; **3**^**rd**^ **wave:** March 21^st^ to July 4^th^ 2021. Results are presented with 95% confidence intervals of the mean total number of contacts and p-values of the difference between individuals with and without comorbidities. *Results adjusted for age **B) Adults over 65 years old** **Pre-COVID:** February 1^st^ 2018 to March 17^th^ 2019; **1**^**st**^ **wave:** April 21^st^ to May 25^th^ 2020; **Summer 2020:** July 3^rd^ to August 22^nd^ 2020; **2**^**nd**^ **wave:** August 23^rd^ 2020 to March 20^th^ 2021; **Holidays:** December 17^th^ 2020 to January 8^th^ 2021; **3**^**rd**^ **wave:** March 21^st^ to July 4^th^ 2021. Results are presented with 95% confidence intervals of the mean total number of contacts and p-values of the difference between individuals with and without comorbidities.

Contacts at home with household members were generally similar for individuals with and without comorbidities and were constant over time (Figure 3, Additional file 1: Table S6B). The differences observed in the total number of social contacts were attributable to contacts with visitors at home and contacts in other locations (Figure 3, Additional file 1: Table S6C). Individuals aged 18 to 65 years with comorbidities had a significantly lower number of contacts in other locations and with visitors at home compared to those without comorbidities in the second wave, excluding the Christmas holidays. For individuals aged over 65 years, the difference was observed during the summer 2020 and the second wave before the Christmas holidays.

### Social contacts in the third wave according to the vaccination status

Vaccination coverage with the first dose of the COVID-19 vaccine increased significantly throughout the third wave, from 28% and 20% at the end of March 2021 for individuals with and without comorbidities, respectively, to 97% and 94% in June-July 2021 (Additional file 1: Figure S2). Considering the entire third wave, vaccination coverage with one dose was similar among individuals with and without comorbidities of all age groups (Additional file 1: Tables S8 and S9).

Vaccination status with the first dose influenced the association between social contacts and comorbidity status (Figure 4, Additional file 1: Table S10). Among unvaccinated individuals, the fewer contacts of those with comorbidities compared to those without comorbidities generally persisted through the third wave. However, among individuals vaccinated with their first dose, there was no significant difference according to the comorbidity status. Moreover, vaccinated individuals had generally more contacts than unvaccinated individuals, irrespective of their comorbidity status. For example, vaccinated individuals with comorbidities aged 26 to 45 years had a significantly higher number of contacts (5.9, 95%CI 3.7– 8.0) than unvaccinated individuals with comorbidities (2.9, 95%CI 1.7–4.1; P=0.02) and reached the level of contacts of vaccinated individuals without comorbidities (6.9, 95%CI 5.4–8.4; P=0.42).

**Figure 4.**
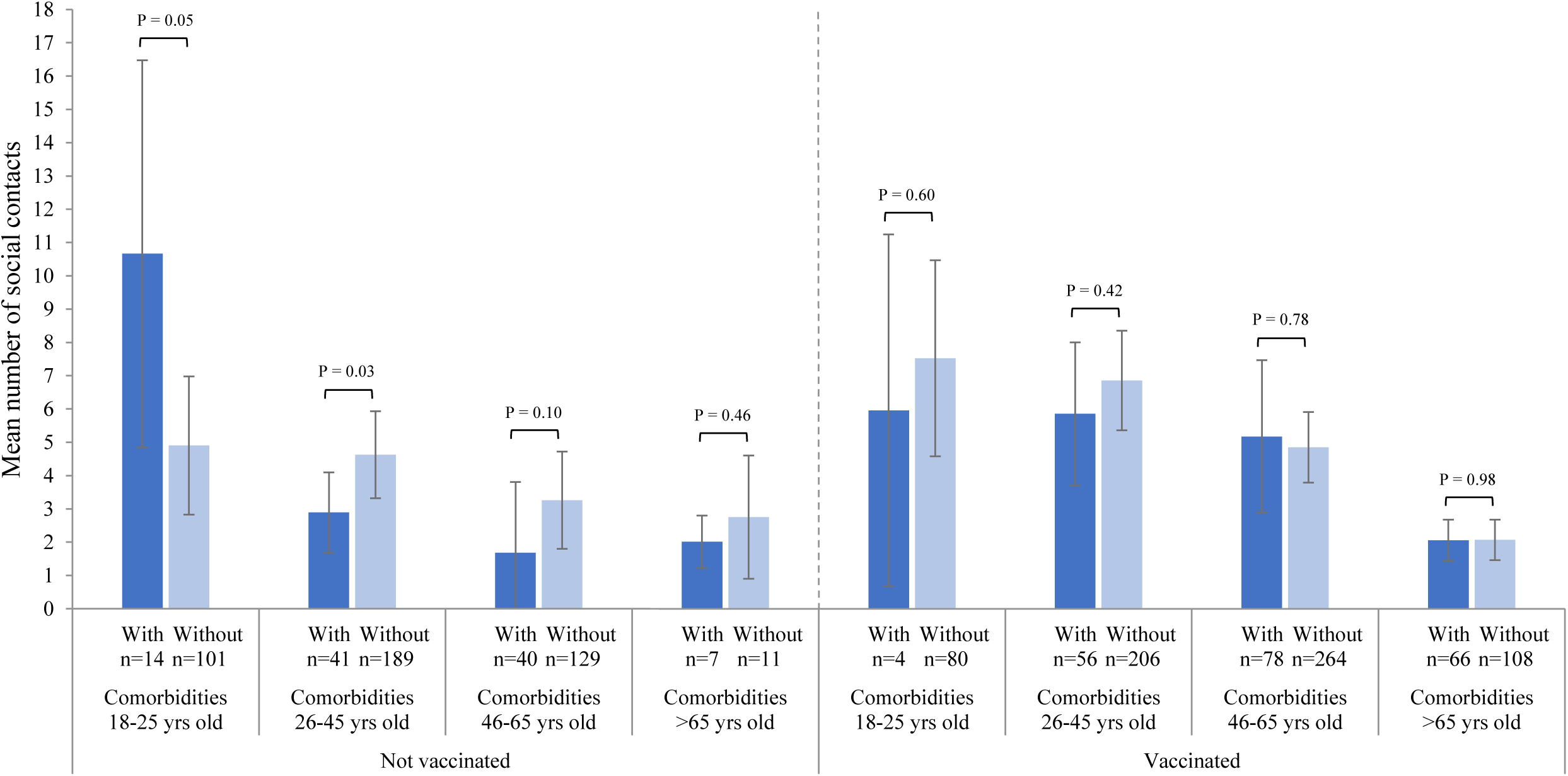
Mean total number of social contacts of individuals with and without comorbidities in the third wave according to vaccination status with one dose. **Third wave:** March 21^st^ to July 4^th^ 2021. Results are adjusted for the time periods (March, April, May, June-July). Results are presented with 95% confidence intervals and p-values of the difference between individuals with and without comorbidities. The “n” in the Figure represent the numbers of unvaccinated and vaccinated adults.

## DISCUSSION

To our knowledge, this is the first study to focus on social contacts of individuals according to comorbidity and COVID-19 vaccination status before and during the COVID-19 pandemic. Our results suggest that individuals with and without comorbidities decreased significantly their contacts during the first wave compared to the pre-pandemic period. During the second wave, individuals with comorbidities maintained fewer social contacts compared to individuals without comorbidities. Individuals aged 18 to 65 years with comorbidities had about 27% fewer social contacts per day than those without comorbidities during the second wave, excluding the Christmas holidays. Similarly, individuals aged over 65 years with comorbidities had 43% fewer social contacts per day than those without comorbidities during the first part of the second wave, before the holidays. Interestingly, as vaccination coverage with the first dose increased in Quebec during the third wave, the differences in the number of contacts between individuals with and without comorbidities declined. During this wave, unvaccinated individuals with comorbidities had generally fewer contacts than those without comorbidities, but there was no significant difference in the total number of social contacts according to the comorbidity status among those vaccinated with their first dose.

Differences in the total number of social contacts according to the comorbidity status during the second wave were explained by differences in contacts with visitors at home and contacts in other locations. Among individuals aged 18 to 65 years, these differences were mainly attributable to contacts at work: 1) fewer individuals with comorbidities were working and 2) working individuals with comorbidities had fewer contacts at work, as recommended for workers with chronic diseases [40]. First, throughout the study periods, significantly fewer participants with comorbidities were employed (62%) compared to participants without comorbidities (72%; P<0.0001). In addition, from summer 2020 to the end of the second wave, there were twice as many participants with comorbidities who were temporarily not working (10%) compared to those without comorbidities (5%, P<0.0001). Second, although their type of employment was similar (Table 1, Additional file 1: Table S5), workers with comorbidities had significantly fewer contacts at work during the second wave (mean 1.9) compared to those without comorbidities (2.8; P=0.02). Among individuals aged over 65 years, few participants were employed, and percentages working were similar among those with (14%) and without comorbidities (15%; P=0.75). The difference seen in the second wave for this age group was mainly attributable to contacts at home with visitors, in leisure activities, and other locations. Of note, there was no difference in contacts according to the comorbidity status during the first wave and the Christmas holidays for all age groups. Contacts were low for all participants during these periods as a result of the complete lockdown during the first wave and school/work breaks and gathering restrictions during the holidays.

Our results support the hypothesis that individuals with comorbidities could have contributed to reducing the severity of the second wave by keeping their contacts at a low level and consequently decreasing their risk of contracting COVID-19. Some studies have hypothesized that a decrease in the proportion of COVID-19 cases with comorbidities could have contributed to the lower severity of the second wave in terms of hospitalisation and death per case compared to the first wave, but sparse data are currently available [7, 41, 42]. The decrease in the severity of the second wave could also be explained by a combination of other factors. First, treatments for COVID-19 improved over time. For example, systemic corticosteroids have been shown to reduce COVID-19 mortality [43], and have been added to the WHO recommendation for the treatment of patients with severe and critical COVID-19 as of September 2020 [44]. Second, screening capacity increased at the end of the first wave. The lower testing capacity during the first wave likely led to an underestimation of the number of cases [42, 45] with a likely higher proportion of more severe cases being detected. Third, non-pharmaceutical interventions, such as physical distancing and use of masks, implemented gradually during the pandemic could have resulted in lower viral inoculum, and some studies suggested that viral load could be associated with COVID-19 disease severity [46, 47].

Our results also support the assumption that vaccination could influence the number of social contacts of individuals, particularly those at greater risk of complications who may feel protected by the vaccine and increase their contacts. During the third wave, the level of contacts of individuals with comorbidities vaccinated with their first dose was similar to those without comorbidities. However, contacts of individuals with comorbidities who were not yet vaccinated when participating in CONNECT remained lower than contacts of unvaccinated individuals without comorbidities. Of note, this observation occurred within the context of the third wave in Quebec, when social contacts increased significantly for all adults compared with the second wave, as vaccination was rolled out and physical distancing measures were gradually relaxed from the end of May 2021. Therefore, our results cannot be extrapolated to currently unvaccinated individuals, as CONNECT participants who were unvaccinated when completing the study questionnaire during the third wave were potentially waiting for their priority group appointment and vaccinated afterwards (Additional file 1: Table S9) [14, 48]. Indeed, vaccination coverage with at least one dose among individuals aged 12 years or older in Quebec exceeded 90% as of October 2021 [49].

Our study has some limitations. First, although CONNECT participants were randomly recruited from the Quebec general population, it is possible that individuals who agreed to participate in the study were more likely to adhere to physical distancing measures aiming to limit social contacts. However, we recruited a large sample of over 6600 participants, and we validated that they were generally representative of the Quebec population in terms of age, sex, region, household composition, race/ethnicity, and vaccination coverage [36, 49]. Second, social desirability bias may have led to underreporting of social contacts. Indeed, because physical distancing measures limited social contacts during the pandemic, some participants may have been reluctant to report all their contacts, especially those not allowed by physical distancing measures. Nevertheless, all questions were identical for participants with and without comorbidities and they remained the same from the pre-pandemic period until the end of the study. Moreover, we ensured that the few added questions related to COVID-19 were asked at the end of the questionnaire to prevent, as much as possible, participants from thinking about COVID-19 and the measures in place when reporting their contacts. Third, comorbidities were self-reported, but we are confident that the most significant comorbidities lasting at least 6 months and for which there were symptoms or medications in the last 12 months were identified. Additionally, the most common comorbidities we identified (chronic lung disease, diabetes, hypertension, chronic heart disease) were similar to those in the Quebec population [6]. These main limitations would likely be non-differential according to the comorbidity status and would likely underestimate the number of contacts of both individuals with and without comorbidities. In addition, it is unclear whether there is a reporting bias of social contacts according to vaccination status. On one hand, if unvaccinated individuals reported fewer contacts than they actually had (only those allowed by physical distancing measures), this might have led to an overestimation of the differences in contacts according to vaccination status. On the other hand, if vaccinated individuals reported fewer contacts than they actually had, while unvaccinated individuals reported all their contacts (whether or not they were allowed by physical distancing measures), we might have an underestimation of the contact differences between vaccinated and unvaccinated individuals. Finally, our exploratory analysis of the influence of vaccination status on the association between contacts and comorbidity status had a limited statistical power as there were few participants in some age-vaccination status categories.

Our study also has five major strengths. First, CONNECT provides pre-pandemic data collected shortly before the pandemic as well as data throughout the first, second, and third waves of pandemic with the same methodology, allowing comparisons of contacts over time. Second, contacts were measured using a validated method that has been used to measure social contacts worldwide for many years [31-33, 50]. Third, our results were robust when using different classifications of comorbidities at risk of COVID-19 complications. Fourth, by collecting various socio-demographic characteristics of the participants, we were able to verify the confounding potential of a large number of variables, thereby limiting confounding bias. Finally, the general idea that individuals with comorbidities could have protected themselves by maintaining a lower level of contacts compared to those without comorbidities and that they may have felt protected by the vaccine and then increased their contacts is likely generalizable to other countries with similar public health measures and COVID-19 vaccine roll-out, as studies from different countries have shown similar decreases in social contacts during the pandemic [1-3]. However, it is important to note that social contacts trends should not be directly interpreted as adherence to public health measures at the individual level. For example, individuals with comorbidities in CONNECT may have felt protected by their first dose of the vaccine and increased their contacts to the same level as those without comorbidities, while still adhering to physical distancing measures.

Our results have important implications. First, they suggest that the lower level of contacts maintained by individuals with comorbidities could have influenced the burden of hospitalisations and deaths related to the different COVID-19 waves. This reduced number of contacts is likely a combination of self-isolation of individuals with comorbidities, who perceived themselves at risk of COVID-19 complications, and efforts of their close contacts to protect them. As shown in other studies and previous CONNECT analyses, individuals who perceive themselves at risk of COVID-19 complications tend to have significantly fewer contacts than those who do not perceive themselves at risk [51-53]. However, keeping extremely low levels of social contacts for several months to decrease the risk of contracting COVID-19 can have a negative impact on mental health. Indeed, several studies have described that social isolation during the pandemic was associated with increased psychological distress, including anxiety and depression [54-56]. Finding ways of mitigating the impact of severely reduced social contacts on mental health would be important when lockdown and physical distancing measures are introduced. Second, our results suggest that individuals at greater risk of complications could have increased their contacts after receiving their first dose of the COVID-19 vaccine, as they may have felt protected by the vaccine. However, in the current context with more transmissible/severe variants with high immune escape, and the waning of two-dose protection, it will be important to closely monitor vaccine efficacy, particularly among populations at high risk of COVID-19 complications who may have returned to higher levels of social contacts. If the vaccine efficacy wanes in these more vulnerable groups, the incremental benefit of booster doses should be examined.

## CONCLUSIONS

In conclusion, the total number of hospitalisations and deaths could have been higher in the second wave in Quebec without the behavior of individuals at greater risk of COVID-19 complications who have maintained a significant reduction of their social contacts. It will be important to closely monitor COVID-19-related outcomes and social contacts by comorbidity and vaccination status to inform targeted or population-based interventions (e.g., booster doses of the vaccine).

## Data Availability

The datasets generated and/or analysed during the current study are not publicly available due to the dataset containing sensitive personal data. Aggregated data are available from the corresponding author upon reasonable request.

## LIST OF ABBREVIATIONS

CI: Confidence interval
INSPQ: Institut national de santé publique du Québec
NACI: National Advisory Committee on Immunization
STROBE: Strengthening the Reporting of Observational Studies in Epidemiology

## DECLARATIONS

### Ethics approval and consent to participate

The CONNECT study was approved by the ethics committee at the Centre de recherche du CHU de Québec-Université Laval (project 2016-2172). All participants provided informed consent during the recruitment phone call.

### Consent for publication

Not applicable.

### Authors’ contributions

MB and MD designed the study, and they developed and validated the questionnaires. MB and MD supervised the data collection and analysis. AG participated in data collection. AG and MM did the statistical analysis. MB, AG and MD drafted the article. AG, MD, MM, MS, CS, GDS and MB interpreted the results and critically revised the manuscript for scientific content. All authors approved the final version of the article.

## Acknowledgements

We thank Éric Demers for managing the database and Norma Pérez for helping with the review of literature.

