## Additional file 1 for "Time trends in social contacts of individuals according to comorbidity and vaccination status, before and during the COVID-19 pandemic"

**Table S1. Distribution of comorbidity status among confirmed cases in Quebec adults***

| | 1st wave | | 2nd wave | | 3rd wave | |
|---|---|---|---|---|---|---|
| | n | % | n | % | n | % |
| **All adults** | | | | | | |
| Without comorbidities | 19265 | 51.2 | 94879 | 56.6 | 29710 | 62.3 |
| With comorbidities | 18331 | 48.8 | 72634 | 43.4 | 17997 | 37.7 |
| **18-45 years old** | | | | | | |
| Without comorbidities | 12529 | 71.0 | 64201 | 73.6 | 20491 | 75.3 |
| With comorbidities | 5119 | 29.0 | 23064 | 26.4 | 6715 | 24.7 |
| **46-65 years old** | | | | | | |
| Without comorbidities | 6169 | 47.3 | 27159 | 50.3 | 8303 | 52.9 |
| With comorbidities | 6877 | 52.7 | 26846 | 49.7 | 7386 | 47.1 |
| **66-75 years old** | | | | | | |
| Without comorbidities | 416 | 17.9 | 2609 | 22.9 | 722 | 25.3 |
| With comorbidities | 1914 | 82.1 | 8787 | 77.1 | 2128 | 74.7 |
| **>75 years old** | | | | | | |
| Without comorbidities | 151 | 3.3 | 910 | 6.1 | 194 | 9.9 |
| With comorbidities | 4421 | 96.7 | 13937 | 93.9 | 1768 | 90.1 |

**1st wave:** February 23rd to July 11th 2020; **2nd wave:** August 23rd 2020 to March 20th 2021; **3rd wave:** March 21st to July 13th 2021.
Methodology used to generate data from the first wave is available online [6]. The same methodology has been used for the other waves.
* Cases from nursing homes are excluded

**Table S2. STROBE Statement—Checklist of items that should be included in reports of *cross-sectional studies* [30]**

| | Item No | Recommendation | Page No |
|---|---|---|---|
| **Title and abstract** | 1 | (*a*) Indicate the study's design with a commonly used term in the title or the abstract | 2 |
| | | (*b*) Provide in the abstract an informative and balanced summary of what was done and what was found | 2, 3 |
| **Introduction** | | | |
| Background/rationale | 2 | Explain the scientific background and rationale for the investigation being reported | 4, 5 |
| Objectives | 3 | State specific objectives, including any prespecified hypotheses | 5, 6 |
| **Methods** | | | |
| Study design | 4 | Present key elements of study design early in the paper | 6 |
| Setting | 5 | Describe the setting, locations, and relevant dates, including periods of recruitment, exposure, follow-up, and data collection | 6, 7, 8, 9 Figure 1 |
| Participants | 6 | (*a*) Give the eligibility criteria, and the sources and methods of selection of participants | 6 |
| Variables | 7 | Clearly define all outcomes, exposures, predictors, potential confounders, and effect modifiers. Give diagnostic criteria, if applicable | 7, 8, 9, 10 |
| Data sources/ measurement | 8* | For each variable of interest, give sources of data and details of methods of assessment (measurement). Describe comparability of assessment methods if there is more than one group | 7, 8, 9 |
| Bias | 9 | Describe any efforts to address potential sources of bias | 9, 10 |
| Study size | 10 | Explain how the study size was arrived at | 10 |
| Quantitative variables | 11 | Explain how quantitative variables were handled in the analyses. If applicable, describe which groupings were chosen and why | 9, 10 |
| Statistical methods | 12 | (*a*) Describe all statistical methods, including those used to control for confounding | 9, 10 |
| | | (*b*) Describe any methods used to examine subgroups and interactions | 9, 10 |
| | | (*c*) Explain how missing data were addressed | 9 |
| | | (*d*) If applicable, describe analytical methods taking account of sampling strategy | N/A |
| | | (*e*) Describe any sensitivity analyses | 10 Appendix p.5 |
| **Results** | | | |
| Participants | 13* | (a) Report numbers of individuals at each stage of study—eg numbers potentially eligible, examined for eligibility, confirmed eligible, included in the study, completing follow-up, and analysed | 10 Appendix p.17 |
| | | (b) Give reasons for non-participation at each stage | 10 |

| | Item No | Recommendation | Page No |
|---|---|---|---|
| | | (c) Consider use of a flow diagram | N/A |
| Descriptive data | 14* | (a) Give characteristics of study participants (eg demographic, clinical, social) and information on exposures and potential confounders | 10, 11 Table 1 Appendix p.7, 8, 17 |
| | | (b) Indicate number of participants with missing data for each variable of interest | 10 |
| Outcome data | 15* | Report numbers of outcome events or summary measures | 11, 12, 13 Figure 2, 3, 4 |
| Main results | 16 | (*a*) Give unadjusted estimates and, if applicable, confounder-adjusted estimates and their precision (eg, 95% confidence interval). Make clear which confounders were adjusted for and why they were included | 11, 12, 13 Appendix p.9, 10, 11 |
| | | (*b*) Report category boundaries when continuous variables were categorized | N/A |
| | | (*c*) If relevant, consider translating estimates of relative risk into absolute risk for a meaningful time period | N/A |
| Other analyses | 17 | Report other analyses done—eg analyses of subgroups and interactions, and sensitivity analyses | 11 Appendix p.12, 13 |
| **Discussion** | | | |
| Key results | 18 | Summarise key results with reference to study objectives | 13 |
| Limitations | 19 | Discuss limitations of the study, taking into account sources of potential bias or imprecision. Discuss both direction and magnitude of any potential bias | 15, 16, 17 |
| Interpretation | 20 | Give a cautious overall interpretation of results considering objectives, limitations, multiplicity of analyses, results from similar studies, and other relevant evidence | 14, 15 |
| Generalisability | 21 | Discuss the generalisability (external validity) of the study results | 17 |
| **Other information** | | | |
| Funding | 22 | Give the source of funding and the role of the funders for the present study and, if applicable, for the original study on which the present article is based | 20 |

*Give information separately for exposed and unexposed groups.

**Table S3. Classifications of active physical comorbidities**

| All active physical comorbidities | Active physical comorbidities at risk of COVID-19 complications according to INSPQ [6] | Active physical comorbidities at risk of COVID-19 complications according to NACI [34] |
|---|---|---|
| Diabetes | Diabetes | Diabetes |
| Chronic heart disease | Chronic heart disease | Chronic heart disease |
| Chronic liver disease | Chronic liver disease | Chronic liver disease |
| Chronic renal disease | Chronic renal disease | Chronic renal disease |
| Alzheimer disease or any other dementia | Alzheimer disease or any other dementia | Alzheimer disease or any other dementia |
| Chronic lung disease | Chronic lung disease | |
| Cancer | Cancer | |
| Immunosuppressing condition | Immunosuppressing condition | |
| Neurologic disease | Neurologic disease | |
| Hematologic disease | Hematologic disease | |
| Thyroid disease | Hypothyroidism | |
| Arrhythmia | Arrhythmia | |
| Vascular disease | Vascular disease | |
| Hypertension | Hypertension | |
| Physical disability | Paralysis | |
| Chronic inflammatory disease | | |
| Asplenia or hyposplenia | | |
| Arthritis or arthrosis | | |
| Dermatological disease | | |
| Multisystemic disease | | |
| Endocrine disease | | |
| Digestive disease | | |
| Lymphatic disease | | |
| Back or spinal condition | | |
| Other chronic pain | | |
| Sensory disability | | |
| Infectious disease | | |
| Sexually transmitted and blood-borne infections | | |
| Orthopedic condition | | |
| Migraine | | |
| Vertigo or Meniere's disease | | |

INSPQ: Institut national de santé publique du Québec
NACI: National Advisory Committee on Immunization

### Table S4. Sample size calculations

#### A) Detectable differences* for different scenarios† of proportion of individuals with comorbidities, sample size, design effect, and mean number of contacts among individuals with comorbidities

| | | Design effect¥ | | | | | | | | | | | | | | | |
|---|---|---|---|---|---|---|---|---|---|---|---|---|---|---|---|---|---|
| | | 1 | | | | 2 | | | | 5 | | | | 10 | | | |
| | | Mean number of contacts among individuals with comorbidities | | | | | | | | | | | | | | | |
| | | 1 | | 3 | | 1 | | 3 | | 1 | | 3 | | 1 | | 3 | |
| % with comorbidities | Sample size | Diff | Power | Diff | Power | Diff | Power | Diff | Power | Diff | Power | Diff | Power | Diff | Power | Diff | Power |
| 22% | 400 | 0.4 | 82 | 0.7 | 87 | 0.6 | 82 | 1.0 | 85 | 1.1 | 83 | 1.6 | 81 | 1.7 | 80 | 2.0 | 68 |
| 22% | 800 | 0.3 | 88 | 0.5 | 89 | 0.4 | 82 | 0.7 | 87 | 0.7 | 83 | 1.1 | 83 | 1.1 | 83 | 1.6 | 81 |
| 22% | 1200 | 0.3 | 97 | 0.4 | 88 | 0.4 | 94 | 0.6 | 90 | 0.6 | 88 | 0.9 | 85 | 0.8 | 80 | 1.3 | 83 |
| 35% | 100 | 0.8 | 86 | 1.2 | 83 | 1.2 | 84 | 1.7 | 80 | 2.0 | 76 | 2.0 | 53 | 2.0 | 47 | 2.0 | 30 |
| 35% | 200 | 0.5 | 83 | 0.8 | 82 | 0.8 | 86 | 1.2 | 83 | 1.3 | 80 | 2.0 | 82 | 2.0 | 76 | 2.0 | 53 |

* Differences of up to 2.0 were considered in power calculations. When a power greater than 80% could not be achieved with the maximum value of 2.0, the power for that difference is presented.

† Scenarios with proportion of 22% with comorbidities and n=400, 800, 1200 represent sample sizes observed for all adults and adults aged 18 to 65 years, and scenarios with proportion of 35% with comorbidities and n=100, 200 represent sample sizes observed for adults aged over 65 years.

¥ The design effect represents the inflation in variance between the observed variance in our study compared to the theoretical variance of a Poisson distribution with simple random sampling.

#### B) Observed design effects for comparisons of contacts between individuals with and without comorbidities*

| | Total contacts | | Contacts at home with household members | | Contacts with visitors at home and contacts in other locations | |
|---|---|---|---|---|---|---|
| | Pre-COVID | During pandemic | Pre-COVID | During pandemic | Pre-COVID | During pandemic |
| All adults | 11.2 | 5.1 to 9.5 | 1.2 | 0.7 to 1.3 | 12.3 | 5.9 to 11.2 |
| 18-65 years old | 19.1 | 7.7 to 12.4 | 1.5 | 0.7 to 1.1 | 20.9 | 9.1 to 15.1 |
| >65 years old | 2.9 | 2.0 to 9.9 | 0.9 | 0.6 to 2.1 | 3.5 | 3.2 to 10.1 |

* Based on comparisons of contacts presented in Table S6.

### Table S5. Key socio-demographic characteristics of participants, by period

| | Pre-COVID | | | | | | 1st wave | | | | | | Summer 2020 and 2nd wave | | | | | | 3rd wave | | | | | |
|---|---|---|---|---|---|---|---|---|---|---|---|---|---|---|---|---|---|---|---|---|---|---|---|---|
| | All participants | | With active physical comorbidities | | Without active physical comorbidities | | All participants | | With active physical comorbidities | | Without active physical comorbidities | | All participants | | With active physical comorbidities | | Without active physical comorbidities | | All participants | | With active physical comorbidities | | Without active physical comorbidities | |
| | $N_{crude}$ | $\%_{weighted}$ | $N_{crude}$ | $\%_{weighted}$ | $N_{crude}$ | $\%_{weighted}$ | $N_{crude}$ | $\%_{weighted}$ | $N_{crude}$ | $\%_{weighted}$ | $N_{crude}$ | $\%_{weighted}$ | $N_{crude}$ | $\%_{weighted}$ | $N_{crude}$ | $\%_{weighted}$ | $N_{crude}$ | $\%_{weighted}$ | $N_{crude}$ | $\%_{weighted}$ | $N_{crude}$ | $\%_{weighted}$ | $N_{crude}$ | $\%_{weighted}$ |
| **Total** | **792** | | **203** | | **589** | | **413** | | **100** | | **313** | | **4125** | | **851** | | **3274** | | **1296** | | **287** | | **1009** | |
| **Age** | | | | | | | | | | | | | | | | | | | | | | | | |
| 18-25 yrs old | 95 | 10.9 | 5 | 1.4 | 90 | 14.1 | 53 | 11.8 | 8 | 5.6 | 45 | 14.2 | 532 | 12.0 | 38 | 3.6 | 494 | 14.2 | 185 | 11.7 | 18 | 4.5 | 167 | 13.8 |
| 26-45 yrs old | 197 | 35.3 | 32 | 26.8 | 165 | 38.2 | 181 | 34.0 | 35 | 20.6 | 146 | 39.1 | 1369 | 34.4 | 193 | 21.6 | 1176 | 37.9 | 457 | 34.2 | 92 | 29.1 | 365 | 35.8 |
| 46-65 yrs old | 294 | 33.0 | 87 | 39.4 | 207 | 30.8 | 129 | 32.8 | 37 | 34.6 | 92 | 32.1 | 1610 | 33.3 | 414 | 39.5 | 1196 | 31.6 | 470 | 32.5 | 106 | 32.2 | 364 | 32.6 |
| 66-75 yrs old | 159 | 12.6 | 60 | 19.2 | 99 | 10.4 | 45 | 12.6 | 16 | 17.0 | 29 | 11.0 | 535 | 12.5 | 167 | 18.1 | 368 | 10.9 | 162 | 12.4 | 62 | 20.6 | 100 | 9.9 |
| >75 yrs old | 47 | 8.3 | 19 | 13.3 | 28 | 6.6 | 5 | 8.8 | 4 | 22.2 | 1 | 3.7 | 79 | 7.9 | 39 | 17.2 | 40 | 5.4 | 22 | 9.3 | 9 | 13.6 | 13 | 8.0 |
| **Sex** | | | | | | | | | | | | | | | | | | | | | | | | |
| Male | 350 | 50.1 | 91 | 49.4 | 259 | 50.3 | 169 | 49.1 | 39 | 49.7 | 130 | 48.9 | 2018 | 49.8 | 413 | 47.8 | 1605 | 50.4 | 577 | 50.2 | 127 | 51.3 | 450 | 49.9 |
| Female | 442 | 49.9 | 112 | 50.6 | 330 | 49.7 | 244 | 50.9 | 61 | 50.3 | 183 | 51.1 | 2107 | 50.2 | 438 | 52.2 | 1669 | 49.6 | 719 | 49.8 | 160 | 48.7 | 559 | 50.2 |
| **Region** | | | | | | | | | | | | | | | | | | | | | | | | |
| Greater Montreal† | 407 | 62.6 | 103 | 62.1 | 304 | 62.7 | 293 | 64.3 | 78 | 67.9 | 215 | 62.9 | 2356 | 61.2 | 486 | 62.8 | 1870 | 60.7 | 749 | 61.1 | 168 | 59.5 | 581 | 61.5 |
| Other Quebec regions | 377 | 37.5 | 99 | 37.9 | 278 | 37.3 | 120 | 35.8 | 22 | 32.1 | 98 | 37.2 | 1766 | 38.8 | 365 | 37.2 | 1401 | 39.3 | 545 | 38.9 | 119 | 40.5 | 426 | 38.5 |
| Missing | 8 | -- | 1 | -- | 7 | -- | -- | -- | -- | -- | -- | -- | 3 | -- | -- | -- | 3 | -- | 2 | -- | -- | -- | 2 | -- |
| **Household size** | | | | | | | | | | | | | | | | | | | | | | | | |
| 1 | 236 | 30.0 | 72 | 35.6 | 164 | 28.2 | 124 | 29.9 | 32 | 28.3 | 92 | 30.5 | 1007 | 24.7 | 252 | 30.5 | 755 | 23.1 | 328 | 27.4 | 77 | 29.3 | 251 | 26.9 |
| 2 | 389 | 43.1 | 107 | 45.9 | 282 | 42.2 | 182 | 45.5 | 53 | 60.5 | 129 | 39.7 | 2086 | 49.3 | 473 | 55.7 | 1613 | 47.5 | 624 | 47.1 | 156 | 52.5 | 468 | 45.5 |
| 3 | 87 | 10.9 | 15 | 10.3 | 72 | 11.1 | 41 | 9.3 | 5 | 3.6 | 36 | 11.5 | 488 | 11.9 | 58 | 6.1 | 430 | 13.5 | 173 | 12.6 | 29 | 9.2 | 144 | 13.7 |
| 4+ | 80 | 16.0 | 9 | 8.2 | 71 | 18.5 | 66 | 15.4 | 10 | 7.6 | 56 | 18.4 | 544 | 14.2 | 68 | 7.7 | 476 | 15.9 | 171 | 12.9 | 25 | 9.1 | 146 | 14.0 |
| **Education Level** | | | | | | | | | | | | | | | | | | | | | | | | |
| No diploma, degree | 64 | 7.8 | 23 | 10.7 | 41 | 6.9 | 20 | 5.1 | 6 | 5.6 | 14 | 4.9 | 172 | 4.1 | 44 | 4.7 | 128 | 3.9 | 45 | 3.5 | 10 | 4.1 | 35 | 3.3 |
| Secondary (high) school | 120 | 14.4 | 28 | 15.1 | 92 | 14.1 | 56 | 14.5 | 14 | 13.6 | 42 | 14.8 | 561 | 13.9 | 118 | 13.8 | 443 | 14.0 | 153 | 11.0 | 32 | 9.8 | 121 | 11.4 |
| College, CEGEP, university or other certificate/diploma | 608 | 77.8 | 152 | 74.2 | 456 | 79.0 | 337 | 80.5 | 80 | 80.8 | 257 | 80.4 | 3392 | 82.0 | 689 | 81.5 | 2703 | 82.1 | 1098 | 85.5 | 245 | 86.2 | 853 | 85.3 |
| **Main occupation** | | | | | | | | | | | | | | | | | | | | | | | | |
| Student employed or unemployed | 76 | 10.8 | 6 | 3.2 | 70 | 13.3 | 51 | 10.4 | 9 | 5.2 | 42 | 12.5 | 481 | 11.1 | 47 | 4.9 | 434 | 12.8 | 153 | 10.3 | 22 | 7.2 | 131 | 11.2 |
| Employed or semi-retired | 363 | 49.5 | 61 | 33.7 | 302 | 54.8 | 230 | 50.6 | 53 | 40.7 | 177 | 54.4 | 2435 | 56.6 | 431 | 45.7 | 2004 | 59.5 | 766 | 55.7 | 143 | 44.4 | 623 | 59.1 |
| Temporarily not working or seeking work | 18 | 2.6 | 6 | 4.0 | 12 | 2.1 | 59 | 12.4 | 12 | 10.3 | 47 | 13.2 | 266 | 5.9 | 75 | 7.5 | 191 | 5.5 | 94 | 6.4 | 25 | 7.5 | 69 | 6.1 |
| Unemployed or retired | 335 | 37.1 | 130 | 59.2 | 205 | 29.8 | 73 | 26.6 | 26 | 43.8 | 47 | 19.9 | 943 | 26.5 | 298 | 41.9 | 645 | 22.2 | 283 | 27.7 | 97 | 40.9 | 186 | 23.7 |

| | Pre-COVID | | | | | | 1st wave | | | | | | Summer 2020 and 2nd wave | | | | | | 3rd wave | | | | | |
|---|---|---|---|---|---|---|---|---|---|---|---|---|---|---|---|---|---|---|---|---|---|---|---|---|
| | All participants | | With active physical comorbidities | | Without active physical comorbidities | | All participants | | With active physical comorbidities | | Without active physical comorbidities | | All participants | | With active physical comorbidities | | Without active physical comorbidities | | All participants | | With active physical comorbidities | | Without active physical comorbidities | |
| | $N_{crude}$ | $\%_{weighted}$ | $N_{crude}$ | $\%_{weighted}$ | $N_{crude}$ | $\%_{weighted}$ | $N_{crude}$ | $\%_{weighted}$ | $N_{crude}$ | $\%_{weighted}$ | $N_{crude}$ | $\%_{weighted}$ | $N_{crude}$ | $\%_{weighted}$ | $N_{crude}$ | $\%_{weighted}$ | $N_{crude}$ | $\%_{weighted}$ | $N_{crude}$ | $\%_{weighted}$ | $N_{crude}$ | $\%_{weighted}$ | $N_{crude}$ | $\%_{weighted}$ |
| **Type of employment*** | | | | | | | | | | | | | | | | | | | | | | | | |
| Education | 54 | 13.5 | 9 | 10.0 | 45 | 14.1 | 27 | 9.1 | 8 | 13.0 | 19 | 7.9 | 228 | 8.5 | 40 | 8.6 | 188 | 8.5 | 90 | 10.0 | 18 | 11.5 | 72 | 9.6 |
| Health | 45 | 11.2 | 9 | 11.2 | 36 | 11.3 | 30 | 9.3 | 6 | 8.1 | 24 | 9.6 | 227 | 8.2 | 46 | 9.4 | 181 | 8.0 | 77 | 8.5 | 18 | 10.5 | 59 | 8.0 |
| Sales and services | 58 | 14.7 | 9 | 18.0 | 49 | 14.0 | 35 | 11.8 | 7 | 11.3 | 28 | 12.0 | 336 | 12.8 | 61 | 13.1 | 275 | 12.7 | 108 | 12.5 | 21 | 13.3 | 87 | 12.3 |
| Other sectors | 215 | 60.6 | 34 | 60.8 | 181 | 60.6 | 188 | 69.8 | 44 | 67.7 | 144 | 70.5 | 1841 | 70.5 | 318 | 68.9 | 1523 | 70.9 | 558 | 69.1 | 94 | 64.8 | 464 | 70.1 |
| Missing | -- | -- | -- | -- | -- | -- | 1 | -- | -- | -- | 1 | -- | 18 | -- | 3 | -- | 15 | -- | 4 | -- | -- | -- | 4 | -- |
| **Race/Ethnicity** | | | | | | | | | | | | | | | | | | | | | | | | |
| White | 728 | 91.6 | 192 | 95.0 | 536 | 90.5 | 355 | 89.3 | 93 | 94.8 | 262 | 87.2 | 3750 | 91.7 | 796 | 95.0 | 2954 | 90.8 | 1150 | 89.9 | 256 | 89.6 | 894 | 90.0 |
| Other | 57 | 8.4 | 11 | 5.1 | 46 | 9.5 | 51 | 10.7 | 7 | 5.2 | 44 | 12.8 | 325 | 8.3 | 44 | 5.0 | 281 | 9.2 | 128 | 10.1 | 26 | 10.4 | 102 | 10.0 |
| Missing | 7 | -- | -- | -- | 7 | -- | 7 | -- | -- | -- | 7 | -- | 50 | -- | 11 | -- | 39 | -- | 18 | -- | 5 | -- | 13 | -- |
| **Country of origin** | | | | | | | | | | | | | | | | | | | | | | | | |
| Canadian-born | 729 | 90.4 | 195 | 94.9 | 534 | 88.9 | 348 | 86.8 | 91 | 94.5 | 257 | 83.9 | 3696 | 88.6 | 789 | 91.9 | 2907 | 87.7 | 1144 | 88.1 | 255 | 87.9 | 889 | 88.2 |
| Foreign-born | 63 | 9.6 | 8 | 5.1 | 55 | 11.1 | 64 | 13.2 | 9 | 5.5 | 55 | 16.1 | 422 | 11.4 | 62 | 8.1 | 360 | 12.3 | 151 | 11.9 | 32 | 12.1 | 119 | 11.8 |
| Missing | -- | -- | -- | -- | -- | -- | 1 | -- | -- | -- | 1 | -- | 7 | -- | -- | -- | 7 | -- | 1 | -- | -- | -- | 1 | -- |
| **Number of active physical comorbidities** | | | | | | | | | | | | | | | | | | | | | | | | |
| 1 | -- | -- | 143 | 71.2 | -- | -- | -- | -- | 79 | 74.6 | -- | -- | -- | -- | 672 | 79.6 | -- | -- | -- | -- | 234 | 82.0 | -- | -- |
| 2 | -- | -- | 44 | 22.2 | -- | -- | -- | -- | 14 | 18.8 | -- | -- | -- | -- | 143 | 15.7 | -- | -- | -- | -- | 45 | 15.6 | -- | -- |
| 3+ | -- | -- | 16 | 6.6 | -- | -- | -- | -- | 7 | 6.5 | -- | -- | -- | -- | 36 | 4.7 | -- | -- | -- | -- | 8 | 2.4 | -- | -- |
| **Type of comorbidities** | | | | | | | | | | | | | | | | | | | | | | | | |
| Chronic lung disease | -- | -- | 41 | 19.4 | -- | -- | -- | -- | 33 | 28.0 | -- | -- | -- | -- | 182 | 20.6 | -- | -- | -- | -- | 59 | 18.6 | -- | -- |
| Diabetes | -- | -- | 54 | 23.7 | -- | -- | -- | -- | 19 | 23.2 | -- | -- | -- | -- | 176 | 20.2 | -- | -- | -- | -- | 59 | 21.6 | -- | -- |
| Chronic inflammatory disease | -- | -- | 35 | 16.7 | -- | -- | -- | -- | 11 | 7.8 | -- | -- | -- | -- | 141 | 17.2 | -- | -- | -- | -- | 49 | 15.0 | -- | -- |
| Hypertension | -- | -- | 25 | 10.7 | -- | -- | -- | -- | 9 | 8.2 | -- | -- | -- | -- | 101 | 14.1 | -- | -- | -- | -- | 30 | 12.5 | -- | -- |
| Chronic heart disease | -- | -- | 29 | 14.6 | -- | -- | -- | -- | 3 | 6.0 | -- | -- | -- | -- | 81 | 9.6 | -- | -- | -- | -- | 24 | 10.2 | -- | -- |
| Cancer | -- | -- | 19 | 9.6 | -- | -- | -- | -- | 8 | 18.9 | -- | -- | -- | -- | 51 | 6.7 | -- | -- | -- | -- | 20 | 7.3 | -- | -- |
| Thyroid disease | -- | -- | 10 | 6.5 | -- | -- | -- | -- | 6 | 4.3 | -- | -- | -- | -- | 60 | 6.9 | -- | -- | -- | -- | 20 | 8.2 | -- | -- |
| Neurologic disease | -- | -- | 14 | 5.4 | -- | -- | -- | -- | 8 | 6.5 | -- | -- | -- | -- | 55 | 5.7 | -- | -- | -- | -- | 12 | 3.9 | -- | -- |
| Arthritis and arthrosis | -- | -- | 16 | 8.3 | -- | -- | -- | -- | 5 | 4.3 | -- | -- | -- | -- | 36 | 4.1 | -- | -- | -- | -- | 8 | 2.5 | -- | -- |
| Others | -- | -- | 36 | 19.5 | -- | -- | -- | -- | 24 | 22.7 | -- | -- | -- | -- | 177 | 19.5 | -- | -- | -- | -- | 66 | 20.4 | -- | -- |

**Pre-COVID:** February 1st 2018 to March 17th 2019; **1st wave:** April 21st to May 25th 2020; **Summer 2020 and 2nd wave:** July 3rd 2020 to March 20th 2021; **3rd wave:** March 21st to July 4th 2021.

† Greater Montreal: Regions of Montreal, Laval, Montérégie, Lanaudière, Laurentides

* Type of employment: among 18-65-year-olds employed or temporarily not working

### Table S6. Time trends in the mean number of social contacts of individuals with and without active physical comorbidities

#### A) Total contacts

| | Pre-COVID n=792 | | 1st wave n=413 | | | Summer 2020 n=679 | | | 2nd wave before holidays n=1261 | | | Holidays n=1197 | | | 2nd wave after holidays n=988 | | | 3rd wave n=1296 | | |
|---|---|---|---|---|---|---|---|---|---|---|---|---|---|---|---|---|---|---|---|---|
| | **mean** | **95% CI** | **mean** | **95% CI** | ***p*[¥]** | **mean** | **95% CI** | ***p*[¥]** | **mean** | **95% CI** | ***p*[¥]** | **mean** | **95% CI** | ***p*[¥]** | **mean** | **95% CI** | ***p*[¥]** | **mean** | **95% CI** | ***p*[¥]** |
| **All adults[*]** **Total** | 7.6 | (6.9-8.3) | 2.9 | (2.5-3.3) | *<.001* | 4.3 | (3.7-4.8) | *<.001* | 3.6 | (3.3-3.9) | *0.05* | 2.9 | (2.6-3.1) | *<.001* | 3.3 | (3.0-3.6) | *0.02* | 4.4 | (4.0-4.7) | *<.001* |
| Without comorbidities | 8.1 | (7.3-9.0) | 2.7 | (2.2-3.2) | *<.001* | 4.3 | (3.6-5.0) | *<.001* | 3.9 | (3.5-4.3) | *0.38* | 2.8 | (2.5-3.1) | *<.001* | 3.4 | (3.0-3.7) | *0.01* | 4.5 | (4.1-4.9) | *<.001* |
| With comorbidities | 6.1 | (4.9-7.3) | 3.2 | (2.5-3.9) | *<.001* | 4.2 | (3.5-4.9) | *0.03* | 2.9 | (2.5-3.2) | *0.001* | 3.0 | (2.6-3.4) | *0.67* | 3.0 | (2.6-3.5) | *0.81* | 4.1 | (3.4-4.7) | *0.009* |
| Diff[†] *(p-value)* | -2.0 | *(0.008)* | 0.5 | *(0.23)* | | -0.1 | *(0.92)* | | -1.0 | *(<.001)* | | 0.2 | *(0.41)* | | -0.3 | *(0.30)* | | -0.4 | *(0.27)* | |
| **18-65 years old[*]** **Total** | 8.9 | (8.0-9.9) | 3.3 | (2.6-3.9) | *<.001* | 4.4 | (3.8-5.0) | *0.009* | 4.2 | (3.8-4.7) | *0.66* | 3.1 | (2.8-3.5) | *<.001* | 3.7 | (3.2-4.1) | *0.05* | 5.1 | (4.6-5.5) | *<.001* |
| Without comorbidities | 9.3 | (8.2-10.4) | 3.2 | (2.5-3.9) | *<.001* | 4.3 | (3.6-5.0) | *0.02* | 4.5 | (4.0-5.0) | *0.66* | 3.1 | (2.7-3.5) | *<.001* | 3.9 | (3.3-4.4) | *0.02* | 5.2 | (4.7-5.7) | *<.001* |
| With comorbidities | 7.7 | (5.7-9.7) | 3.5 | (1.9-5.0) | *0.001* | 4.9 | (3.6-6.3) | *0.16* | 3.2 | (2.6-3.8) | *0.03* | 3.2 | (2.4-4.0) | *0.91* | 2.8 | (2.1-3.6) | *0.51* | 4.6 | (3.6-5.6) | *0.006* |
| Diff[†] *(p-value)* | -1.6 | *(0.16)* | 0.3 | *(0.76)* | | 0.7 | *(0.40)* | | -1.2 | *(0.003)* | | 0.1 | *(0.84)* | | -1.0 | *(0.03)* | | -0.6 | *(0.34)* | |
| **>65 years old** **Total** | 3.5 | (3.0-4.0) | 1.2 | (0.7-1.6) | *<.001* | 2.8 | (1.9-3.7) | *0.002* | 1.5 | (1.3-1.8) | *0.008* | 1.2 | (1.0-1.5) | *0.10* | 1.5 | (1.2-1.9) | *0.13* | 2.1 | (1.7-2.6) | *0.05* |
| Without comorbidities | 4.0 | (3.3-4.8) | 1.0 | (0.3-1.7) | *<.001* | 3.3 | (1.8-4.7) | *0.006* | 1.8 | (1.5-2.0) | *0.05* | 1.3 | (0.9-1.6) | *0.01* | 1.5 | (1.1-2.0) | *0.35* | 2.1 | (1.5-2.8) | *0.12* |
| With comorbidities | 2.8 | (2.3-3.3) | 1.3 | (0.8-1.8) | *<.001* | 2.1 | (1.4-2.7) | *0.07* | 1.0 | (0.6-1.4) | *0.009* | 1.2 | (0.9-1.5) | *0.46* | 1.6 | (1.1-2.1) | *0.18* | 2.1 | (1.4-2.8) | *0.22* |
| Diff[†] *(p-value)* | -1.2 | *(0.006)* | 0.3 | *(0.49)* | | -1.2 | *(0.13)* | | -0.8 | *(<.001)* | | -0.1 | *(0.78)* | | 0.1 | *(0.87)* | | -0.0 | *(0.93)* | |

**Pre-COVID:** February 1st 2018 to March 17th 2019; **1st wave:** April 21st to May 25th 2020; **Summer 2020:** July 3rd to August 22nd 2020; **2nd wave:** August 23rd 2020 to March 20th 2021; **Holidays:** December 17th 2020 to January 8th 2021; **3rd wave:** March 21st to July 4th 2021.

[¥] p: p-value of the difference with the preceding period

[*] Results for all adults and 18-65-year-olds are adjusted for age

[†] Diff: Difference between individuals with and without comorbidities and p-value of this difference

### B) Contacts at home with household members

| | Pre-COVID n=792 | | 1st wave n=413 | | | Summer 2020 n=679 | | | 2nd wave before holidays n=1261 | | | Holidays n=1197 | | | 2nd wave after holidays n=988 | | | 3rd wave n=1296 | | |
|---|---|---|---|---|---|---|---|---|---|---|---|---|---|---|---|---|---|---|---|---|
| | **mean** | **95% CI** | **mean** | **95% CI** | ***p*[¥]** | **mean** | **95% CI** | ***p*[¥]** | **mean** | **95% CI** | ***p*[¥]** | **mean** | **95% CI** | ***p*[¥]** | **mean** | **95% CI** | ***p*[¥]** | **mean** | **95% CI** | ***p*[¥]** |
| **All adults[*]** **Total** | 0.9 | (0.8-1.0) | 0.9 | (0.8-1.1) | *0.33* | 0.9 | (0.8-1.0) | *0.80* | 1.0 | (0.9-1.0) | *0.21* | 1.0 | (0.9-1.0) | *0.97* | 1.0 | (1.0-1.1) | *0.26* | 1.0 | (0.9-1.0) | *0.15* |
| Without comorbidities | 0.9 | (0.8-1.0) | 0.9 | (0.8-1.0) | *0.43* | 0.9 | (0.8-1.0) | *0.80* | 1.0 | (1.0-1.1) | *0.04* | 1.0 | (0.9-1.1) | *0.49* | 1.0 | (1.0-1.1) | *0.29* | 1.0 | (0.9-1.0) | *0.11* |
| With comorbidities | 0.9 | (0.8-1.0) | 1.0 | (0.7-1.2) | *0.57* | 1.0 | (0.8-1.1) | *0.99* | 0.9 | (0.8-1.0) | *0.31* | 1.0 | (0.9-1.1) | *0.25* | 1.0 | (0.9-1.1) | *0.79* | 1.0 | (0.9-1.1) | *0.98* |
| Diff[†] *(p-value)* | 0.0 | *(0.61)* | 0.1 | *(0.69)* | | 0.1 | *(0.42)* | | -0.1 | *(0.03)* | | -0.0 | *(0.70)* | | -0.1 | *(0.40)* | | 0.0 | *(0.72)* | |
| **18-65 years old[*]** **Total** | 1.0 | (0.9-1.1) | 1.0 | (0.9-1.1) | *0.63* | 1.0 | (0.9-1.1) | *0.71* | 1.1 | (1.0-1.1) | *0.10* | 1.1 | (1.0-1.2) | *0.87* | 1.1 | (1.1-1.2) | *0.36* | 1.1 | (1.1-1.2) | *0.58* |
| Without comorbidities | 1.0 | (0.9-1.1) | 1.1 | (1.0-1.2) | *0.24* | 1.0 | (0.9-1.1) | *0.23* | 1.1 | (1.0-1.2) | *0.08* | 1.1 | (1.0-1.2) | *0.93* | 1.2 | (1.1-1.2) | *0.30* | 1.1 | (1.0-1.2) | *0.30* |
| With comorbidities | 1.0 | (0.8-1.2) | 0.8 | (0.7-1.0) | *0.13* | 1.0 | (0.9-1.2) | *0.09* | 1.0 | (0.9-1.2) | *0.83* | 1.1 | (0.9-1.2) | *0.90* | 1.0 | (0.9-1.2) | *0.81* | 1.1 | (1.0-1.3) | *0.31* |
| Diff[†] *(p-value)* | 0.0 | *(0.96)* | -0.3 | *(0.008)* | | 0.0 | *(0.83)* | | -0.1 | *(0.38)* | | -0.1 | *(0.50)* | | -0.1 | *(0.09)* | | 0.0 | *(0.76)* | |
| **>65 years old** **Total** | 0.4 | (0.3-0.5) | 0.6 | (0.3-0.8) | *0.37* | 0.5 | (0.4-0.7) | *0.87* | 0.6 | (0.5-0.7) | *0.69* | 0.6 | (0.5-0.7) | *0.86* | 0.6 | (0.5-0.7) | *0.48* | 0.5 | (0.4-0.6) | *0.08* |
| Without comorbidities | 0.4 | (0.3-0.5) | 0.3 | (0.1-0.5) | *0.58* | 0.5 | (0.3-0.8) | *0.25* | 0.7 | (0.6-0.8) | *0.21* | 0.6 | (0.5-0.7) | *0.20* | 0.6 | (0.5-0.7) | *0.78* | 0.5 | (0.3-0.6) | *0.18* |
| With comorbidities | 0.5 | (0.4-0.6) | 0.8 | (0.4-1.2) | *0.15* | 0.6 | (0.4-0.8) | *0.34* | 0.4 | (0.2-0.6) | *0.22* | 0.6 | (0.4-0.7) | *0.15* | 0.6 | (0.5-0.8) | *0.48* | 0.5 | (0.4-0.7) | *0.28* |
| Diff[†] *(p-value)* | 0.1 | *(0.32)* | 0.5 | *(0.04)* | | 0.1 | *(0.71)* | | -0.3 | *(0.01)* | | -0.0 | *(0.99)* | | 0.0 | *(0.65)* | | 0.1 | *(0.59)* | |

**Pre-COVID:** February 1st 2018 to March 17th 2019; **1st wave:** April 21st to May 25th 2020; **Summer 2020:** July 3rd to August 22nd 2020; **2nd wave:** August 23rd 2020 to March 20th 2021; **Holidays:** December 17th 2020 to January 8th 2021; **3rd wave:** March 21st to July 4th 2021.

[¥] p: p-value of the difference with the preceding period

[*] Results for all adults and 18-65-year-olds are adjusted for age

[†] Diff: Difference between individuals with and without comorbidities and p-value of this difference

### C) Contacts with visitors at home and contacts in other locations

| | Pre-COVID n=792 | | 1st wave n=413 | | | Summer 2020 n=679 | | | 2nd wave before holidays n=1261 | | | Holidays n=1197 | | | 2nd wave after holidays n=988 | | | 3rd wave n=1296 | | |
|---|---|---|---|---|---|---|---|---|---|---|---|---|---|---|---|---|---|---|---|---|
| | **mean** | **95% CI** | **mean** | **95% CI** | ***p*[¥]** | **mean** | **95% CI** | ***p*[¥]** | **mean** | **95% CI** | ***p*[¥]** | **mean** | **95% CI** | ***p*[¥]** | **mean** | **95% CI** | ***p*[¥]** | **mean** | **95% CI** | ***p*[¥]** |
| **All adults[*]** **Total** | 6.8 | (6.1-7.5) | 2.0 | (1.7-2.3) | *<.001* | 3.3 | (2.8-3.8) | *<.001* | 2.6 | (2.3-2.9) | *0.02* | 1.9 | (1.7-2.1) | *<.001* | 2.2 | (2.0-2.5) | *0.05* | 3.4 | (3.1-3.8) | *<.001* |
| Without comorbidities | 7.3 | (6.4-8.2) | 1.8 | (1.4-2.2) | *<.001* | 3.4 | (2.7-4.0) | *<.001* | 2.8 | (2.5-3.2) | *0.17* | 1.8 | (1.6-2.1) | *<.001* | 2.3 | (1.9-2.6) | *0.03* | 3.5 | (3.1-3.9) | *<.001* |
| With comorbidities | 5.3 | (4.0-6.5) | 2.1 | (1.6-2.6) | *<.001* | 3.2 | (2.5-3.8) | *0.009* | 2.0 | (1.7-2.3) | *0.002* | 2.0 | (1.6-2.3) | *0.95* | 2.1 | (1.7-2.5) | *0.74* | 3.1 | (2.5-3.7) | *0.005* |
| Diff[†] *(p-value)* | -2.1 | *(0.008)* | 0.3 | *(0.29)* | | -0.2 | *(0.68)* | | -0.8 | *(<.001)* | | 0.2 | *(0.44)* | | -0.2 | *(0.40)* | | -0.5 | *(0.23)* | |
| **18-65 years old[*]** **Total** | 8.0 | (7.0-8.9) | 2.2 | (1.6-2.8) | *<.001* | 3.4 | (2.8-4.0) | *0.006* | 3.1 | (2.7-3.6) | *0.42* | 2.0 | (1.7-2.4) | *<.001* | 2.5 | (2.1-3.0) | *0.06* | 4.0 | (3.5-4.4) | *<.001* |
| Without comorbidities | 8.3 | (7.2-9.4) | 2.1 | (1.4-2.8) | *<.001* | 3.3 | (2.7-4.0) | *0.01* | 3.3 | (2.8-3.8) | *0.99* | 2.0 | (1.6-2.4) | *<.001* | 2.7 | (2.2-3.2) | *0.03* | 4.1 | (3.5-4.6) | *<.001* |
| With comorbidities | 6.7 | (4.7-8.7) | 2.7 | (1.2-4.2) | *0.001* | 3.9 | (2.5-5.3) | *0.23* | 2.2 | (1.6-2.8) | *0.03* | 2.1 | (1.3-2.9) | *0.89* | 1.8 | (1.1-2.6) | *0.56* | 3.5 | (2.5-4.5) | *0.007* |
| Diff[†] *(p-value)* | -1.6 | *(0.17)* | 0.6 | *(0.51)* | | 0.6 | *(0.47)* | | -1.1 | *(0.004)* | | 0.2 | *(0.74)* | | -0.9 | *(0.06)* | | -0.6 | *(0.33)* | |
| **>65 years old** **Total** | 3.1 | (2.6-3.6) | 0.6 | (0.3-0.9) | *<.001* | 2.2 | (1.4-3.0) | *<.001* | 0.9 | (0.7-1.2) | *0.003* | 0.7 | (0.4-0.9) | *0.09* | 0.9 | (0.6-1.2) | *0.17* | 1.6 | (1.2-2.1) | *0.01* |
| Without comorbidities | 3.6 | (2.9-4.4) | 0.7 | (0.1-1.2) | *<.001* | 2.8 | (1.5-4.0) | *0.003* | 1.1 | (0.8-1.4) | *0.01* | 0.7 | (0.4-1.0) | *0.05* | 0.9 | (0.5-1.4) | *0.37* | 1.7 | (1.1-2.3) | *0.05* |
| With comorbidities | 2.3 | (1.8-2.8) | 0.5 | (0.3-0.8) | *<.001* | 1.5 | (0.8-2.1) | *0.006* | 0.6 | (0.3-0.9) | *0.02* | 0.6 | (0.4-0.9) | *0.98* | 0.9 | (0.5-1.4) | *0.22* | 1.6 | (0.9-2.2) | *0.11* |
| Diff[†] *(p-value)* | -1.3 | *(0.004)* | -0.2 | *(0.59)* | | -1.3 | *(0.07)* | | -0.5 | *(0.02)* | | -0.1 | *(0.76)* | | 0.0 | *(0.97)* | | -0.1 | *(0.83)* | |

**Pre-COVID:** February 1st 2018 to March 17th 2019; **1st wave:** April 21st to May 25th 2020; **Summer 2020:** July 3rd to August 22nd 2020; **2nd wave:** August 23rd 2020 to March 20th 2021; **Holidays:** December 17th 2020 to January 8th 2021; **3rd wave:** March 21st to July 4th 2021.

[¥] p: p-value of the difference with the preceding period

[*] Results for all adults and 18-65-year-olds are adjusted for age

[†] Diff: Difference between individuals with and without comorbidities and p-value of this difference

### Table S7. Time trends in the mean total number of social contacts of individuals with and without active physical comorbidities at risk of COVID-19 complications

### A) According to INSPQ

| | Pre-COVID n=792 | | 1st wave n=413 | | | Summer 2020 n=679 | | | 2nd wave before holidays n=1261 | | | Holidays n=1197 | | | 2nd wave after holidays n=988 | | | 3rd wave n=1296 | | |
|---|---|---|---|---|---|---|---|---|---|---|---|---|---|---|---|---|---|---|---|---|
| | **mean** | **95% CI** | **mean** | **95% CI** | ***p*[¥]** | **mean** | **95% CI** | ***p*[¥]** | **mean** | **95% CI** | ***p*[¥]** | **mean** | **95% CI** | ***p*[¥]** | **mean** | **95% CI** | ***p*[¥]** | **mean** | **95% CI** | ***p*[¥]** |
| **All adults[*]** | | | | | | | | | | | | | | | | | | | | |
| Without comorbidities | 7.9 | (7.1-8.7) | 2.8 | (2.3-3.3) | *<.001* | 4.4 | (3.7-5.0) | *<.001* | 3.8 | (3.4-4.1) | *0.12* | 2.9 | (2.6-3.1) | *<.001* | 3.3 | (3.0-3.7) | *0.04* | 4.4 | (4.0-4.8) | *<.001* |
| With comorbidities | 6.3 | (4.7-7.8) | 3.0 | (2.4-3.7) | *<.001* | 3.7 | (3.1-4.3) | *0.11* | 3.0 | (2.6-3.4) | *0.05* | 2.8 | (2.4-3.1) | *0.44* | 3.1 | (2.6-3.7) | *0.29* | 4.1 | (3.4-4.9) | *0.02* |
| Diff[†] *(p-value)* | -1.6 | *(0.06)* | 0.2 | *(0.68)* | | -0.7 | *(0.14)* | | -0.8 | *(0.002)* | | -0.1 | *(0.74)* | | -0.2 | *(0.55)* | | -0.3 | *(0.47)* | |
| **18-65 years old[*]** | | | | | | | | | | | | | | | | | | | | |
| Without comorbidities | 9.1 | (8.1-10.1) | 3.4 | (2.7-4.1) | *<.001* | 4.5 | (3.8-5.2) | *0.03* | 4.4 | (3.9-4.9) | *0.79* | 3.2 | (2.8-3.6) | *<.001* | 3.8 | (3.3-4.3) | *0.07* | 5.1 | (4.6-5.6) | *<.001* |
| With comorbidities | 8.0 | (5.3-10.7) | 2.6 | (1.1-4.1) | *0.001* | 3.9 | (3.0-4.9) | *0.13* | 3.3 | (2.6-4.1) | *0.32* | 2.6 | (1.9-3.2) | *0.10* | 2.9 | (2.0-3.9) | *0.51* | 4.8 | (3.5-6.1) | *0.02* |
| Diff[†] *(p-value)* | -1.0 | *(0.48)* | -0.8 | *(0.32)* | | -0.6 | *(0.35)* | | -1.0 | *(0.02)* | | -0.6 | *(0.09)* | | -0.8 | *(0.12)* | | -0.3 | *(0.68)* | |
| **>65 years old** | | | | | | | | | | | | | | | | | | | | |
| Without comorbidities | 3.8 | (3.2-4.5) | 1.0 | (0.3-1.7) | *<.001* | 3.2 | (2.0-4.5) | *0.003* | 1.7 | (1.4-1.9) | *0.02* | 1.3 | (1.0-1.6) | *0.06* | 1.5 | (1.1-1.9) | *0.33* | 2.1 | (1.5-2.8) | *0.10* |
| With comorbidities | 2.9 | (2.4-3.4) | 1.3 | (0.8-1.8) | *<.001* | 1.8 | (1.1-2.5) | *0.28* | 1.1 | (0.7-1.5) | *0.11* | 1.2 | (0.8-1.5) | *0.86* | 1.6 | (1.0-2.2) | *0.19* | 2.1 | (1.4-2.8) | *0.30* |
| Diff[†] *(p-value)* | -1.0 | *(0.02)* | 0.3 | *(0.47)* | | -1.4 | *(0.05)* | | -0.6 | *(0.03)* | | -0.1 | *(0.65)* | | 0.1 | *(0.84)* | | -0.1 | *(0.90)* | |

INSPQ: Institut national de santé publique du Québec

**Pre-COVID:** February 1st 2018 to March 17th 2019; **1st wave:** April 21st to May 25th 2020; **Summer 2020:** July 3rd to August 22nd 2020; **2nd wave:** August 23rd 2020 to March 20th 2021; **Holidays:** December 17th 2020 to January 8th 2021; **3rd wave:** March 21st to July 4th 2021.

[¥] p: p-value of the difference with the preceding period

[*] Results for all adults and 18-65-year-olds are adjusted for age

[†] Diff: Difference between individuals with and without comorbidities and p-value of this difference

### B) According to NACI

| | Pre-COVID n=792 | | 1st wave n=413 | | | Summer 2020 n=679 | | | 2nd wave before holidays n=1261 | | | Holidays n=1197 | | | 2nd wave after holidays n=988 | | | 3rd wave n=1296 | | |
|---|---|---|---|---|---|---|---|---|---|---|---|---|---|---|---|---|---|---|---|---|
| | **mean** | **95% CI** | **mean** | **95% CI** | ***p*[¥]** | **mean** | **95% CI** | ***p*[¥]** | **mean** | **95% CI** | ***p*[¥]** | **mean** | **95% CI** | ***p*[¥]** | **mean** | **95% CI** | ***p*[¥]** | **mean** | **95% CI** | ***p*[¥]** |
| **All adults[*]** | | | | | | | | | | | | | | | | | | | | |
| Without comorbidities | 7.9 | (7.1-8.7) | 2.9 | (2.5-3.4) | *<.001* | 4.3 | (3.8-4.9) | *<.001* | 3.7 | (3.4-4.0) | *0.06* | 2.8 | (2.6-3.1) | *<.001* | 3.3 | (3.0-3.6) | *0.01* | 4.4 | (4.0-4.8) | *<.001* |
| With comorbidities | 5.1 | (4.1-6.1) | 2.7 | (2.2-3.2) | *<.001* | 3.1 | (2.4-3.7) | *0.41* | 2.8 | (2.3-3.2) | *0.46* | 3.0 | (2.5-3.4) | *0.55* | 2.8 | (2.1-3.5) | *0.75* | 4.1 | (3.0-5.1) | *0.05* |
| Diff[†] *(p-value)* | -2.8 | *(<.001)* | -0.2 | *(0.52)* | | -1.3 | *(0.004)* | | -0.9 | *(0.001)* | | 0.1 | *(0.61)* | | -0.5 | *(0.19)* | | -0.4 | *(0.54)* | |
| **18-65 years old[*]** | | | | | | | | | | | | | | | | | | | | |
| Without comorbidities | 9.1 | (8.1-10.1) | 3.4 | (2.7-4.0) | *<.001* | 4.5 | (3.8-5.1) | *0.02* | 4.3 | (3.9-4.8) | *0.72* | 3.1 | (2.8-3.5) | *<.001* | 3.8 | (3.3-4.2) | *0.03* | 5.1 | (4.6-5.5) | *<.001* |
| With comorbidities | 5.9 | (3.8-8.1) | 1.2 | (0.9-1.6) | *<.001* | 3.4 | (2.0-4.9) | *0.003* | 2.6 | (1.9-3.4) | *0.33* | 2.8 | (2.0-3.7) | *0.75* | 2.0 | (1.5-2.6) | *0.12* | 5.3 | (2.6-8.0) | *0.02* |
| Diff[†] *(p-value)* | -3.2 | *(0.008)* | -2.1 | *(<.001)* | | -1.0 | *(0.19)* | | -1.7 | *(<.001)* | | -0.3 | *(0.50)* | | -1.7 | *(<.001)* | | 0.3 | *(0.85)* | |
| **>65 years old** | | | | | | | | | | | | | | | | | | | | |
| Without comorbidities | 3.7 | (3.1-4.3) | 1.2 | (0.7-1.8) | *<.001* | 3.0 | (2.0-4.0) | *0.003* | 1.5 | (1.3-1.8) | *0.007* | 1.3 | (1.0-1.5) | *0.16* | 1.6 | (1.2-2.0) | *0.22* | 2.2 | (1.7-2.8) | *0.06* |
| With comorbidities | 3.0 | (2.3-3.6) | 1.0 | (0.4-1.7) | *<.001* | 1.4 | (0.5-2.3) | *0.52* | 1.3 | (0.6-2.0) | *0.86* | 1.1 | (0.7-1.5) | *0.62* | 1.4 | (1.0-1.9) | *0.32* | 1.7 | (1.0-2.4) | *0.45* |
| Diff[†] *(p-value)* | -0.7 | *(0.10)* | -0.2 | *(0.69)* | | -1.6 | *(0.02)* | | -0.2 | *(0.53)* | | -0.2 | *(0.52)* | | -0.2 | *(0.62)* | | -0.5 | *(0.30)* | |

NACI: National Advisory Committee on Immunization

**Pre-COVID:** February 1st 2018 to March 17th 2019; **1st wave:** April 21st to May 25th 2020; **Summer 2020:** July 3rd to August 22nd 2020; **2nd wave:** August 23rd 2020 to March 20th 2021; **Holidays:** December 17th 2020 to January 8th 2021; **3rd wave:** March 21st to July 4th 2021.

[¥] p: p-value of the difference with the preceding period

[*] Results for all adults and 18-65-year-olds are adjusted for age

[†] Diff: Difference between individuals with and without comorbidities and p-value of this difference

### Table S8. Vaccination coverage with one dose of individuals with and without active physical comorbidities in the third wave

| | 18-25 yrs old n=199 | | 26-45 yrs old n=492 | | 46-65 yrs old n=511 | | >65 yrs old n=192 | |
|---|---|---|---|---|---|---|---|---|
| | **mean (%)** | **95% CI** | **mean (%)** | **95% CI** | **mean (%)** | **95% CI** | **mean (%)** | **95% CI** |
| Without comorbidities | 44.0 | (36.6-51.4) | 52.0 | (46.7-57.1) | 67.6 | (62.9-72.3) | 89.1 | (80.1-98.0) |
| With comorbidities | 25.4 | (3.8-47.0) | 57.9 | (47.7-68.2) | 66.5 | (57.7-75.2) | 93.8 | (89.0-98.5) |
| Diff† *(p-value)* | -18.6 | *(0.11)* | 6.0 | *(0.31)* | -1.1 | *(0.83)* | 4.7 | *(0.36)* |

**Third wave:** March 21st to July 4th 2021.

† Diff: Difference between individuals with and without comorbidities and p-value of this difference

**Table S9. Recommended Quebec priority groups for vaccination against COVID-19 [14]**

| Priority order | Groups |
|---|---|
| **1** | Elderly people living in long-term care facilities (nursing homes, CHSLD) |
| **2** | Health care workers |
| **3** | Elderly people living in retirement homes |
| **4** | Isolated and remote communities |
| **5** | Adults aged over 80 years |
| **6** | Adults aged 70 to 79 years |
| **7** | Adults aged 60 to 69 years |
| **8** | Adults aged under 60 years with comorbidities at risk of COVID-19 complications* |
| **9** | Adults aged under 60 years without comorbidities working in essential services |
| **10** | General adult population |
| **11** | Children |

* In practice, only individuals aged under 60 years with conditions at very high risk of COVID-19 complications (hospitalised patients with comorbidities or outpatients with severe immunosuppressive conditions) were invited to be vaccinated before other individuals in their age group [48].

**Table S10. Mean total number of social contacts in the third wave of individuals with and without active physical comorbidities according to vaccination status with one dose**

| | 18-25 yrs old n=199 | | | 26-45 yrs old n=492 | | | 46-65 yrs old n=511 | | | >65 yrs old n=192 | | |
|---|---|---|---|---|---|---|---|---|---|---|---|---|
| | **n** | **mean** | **95% CI** | **n** | **mean** | **95% CI** | **n** | **mean** | **95% CI** | **n** | **mean** | **95% CI** |
| **Not vaccinated** | | | | | | | | | | | | |
| Without comorbidities | 101 | 4.9 | (2.8-7.0) | 189 | 4.6 | (3.3-5.9) | 129 | 3.3 | (1.8-4.7) | 11 | 2.8 | (0.9-4.6) |
| With comorbidities | 14 | 10.7 | (4.9-16.5) | 41 | 2.9 | (1.7-4.1) | 40 | 1.7 | (0.0-3.8) | 7 | 2.0 | (1.2-2.8) |
| **Vaccinated** | | | | | | | | | | | | |
| Without comorbidities | 80 | 7.5 | (4.6-10.5) | 206 | 6.9 | (5.4-8.4) | 264 | 4.8 | (3.8-5.9) | 108 | 2.1 | (1.5-2.7) |
| With comorbidities | 4 | 6.0 | (0.7-11.2) | 56 | 5.9 | (3.7-8.0) | 78 | 5.2 | (2.9-7.5) | 66 | 2.1 | (1.4-2.7) |
| **Between group differences** | | | | | | | | | | | | |
| **Not vaccinated:** with – without comorbidities *(p-value)* | | 5.8 | *(0.05)* | | -1.7 | *(0.03)* | | -1.6 | *(0.10)* | | -0.7 | *(0.46)* |
| **Vaccinated:** with – without comorbidities *(p-value)* | | -1.6 | *(0.60)* | | -1.0 | *(0.42)* | | 0.3 | *(0.78)* | | -0.0 | *(0.98)* |
| **Without comorbidities:** vaccinated – not vaccinated *(p-value)* | | 2.6 | *(0.21)* | | 2.2 | *(0.05)* | | 1.6 | *(0.13)* | | -0.7 | *(0.47)* |
| **With comorbidities:** vaccinated – not vaccinated *(p-value)* | | -4.7 | *(0.25)* | | 3.0 | *(0.02)* | | 3.5 | *(0.06)* | | 0.1 | *(0.92)* |

**Third wave:** March 21st to July 4th 2021.
Results are adjusted for the time periods (March, April, May, June-July).

**Figure S1. Distribution of participants according to classifications of comorbidities**

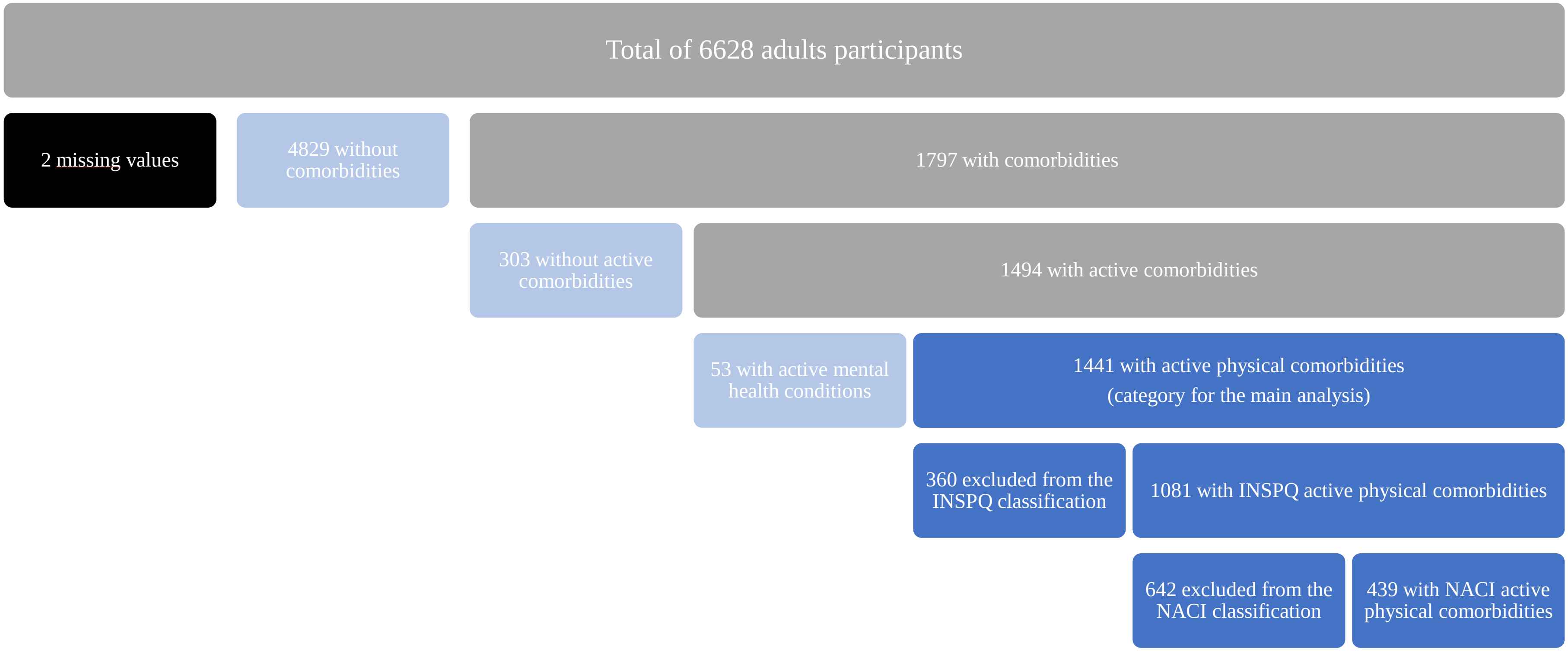

INSPQ: Institut national de santé publique du Québec
NACI: National Advisory Committee on Immunization

**Figure S2. Time trends in vaccination coverage with one dose of individuals with and without active physical comorbidities in the third wave**

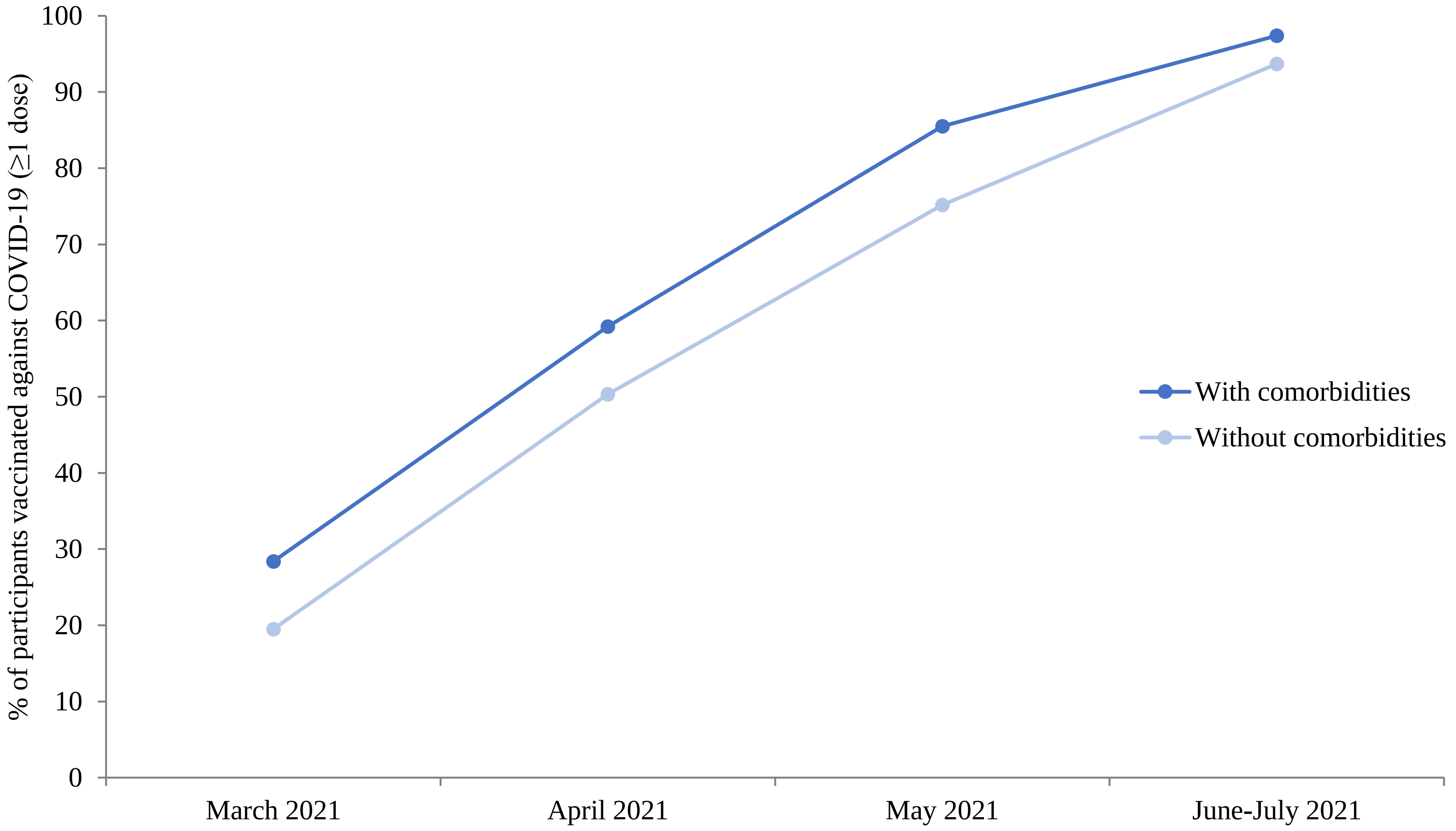

**Third wave:** March 21st to July 4th 2021.

**Example of questions S1. Questions on health conditions**

**27. A) Which of the following long-term conditions do you have or have had?**
***(check all that apply)***

**Health conditions**

- ☐ Chronic lung disease (e.g., asthma, chronic bronchitis, emphysema, chronic obstructive pulmonary disease (COPD), cystic fibrosis)
- ☐ Diabetes
- ☐ Chronic heart disease
- ☐ Chronic liver disease
- ☐ Chronic inflammatory disease (e.g., lupus erythematosus, rheumatoid arthritis, psoriasis, eczema, Crohn's disease, ulcerative colitis)
- ☐ Cancer
- ☐ Immunosuppressing condition (e.g., organ transplantation, HIV-infection, cancer chemotherapy, radiotherapy, congenital immunodeficiency)
- ☐ Chronic renal disease and/or dialysis
- ☐ Asplenia or hyposplenia (absent or defective spleen)
- ☐ Neurologic problem (e.g., Guillain-Barré syndrome, multiple sclerosis)
- ☐ Hematologic problem (e.g., anemia, hemoglobinopathy, bleeding problems)
- ☐ Alzheimer disease or any other dementia
- ☐ Another long-term health condition, specify: ____________________

### 27. B] For which of the following conditions have you had any symptoms or taken any medicine **<u>in the past 12 months</u>** ?

**Health conditions**

- ☐ Chronic lung disease (e.g., asthma, chronic bronchitis, emphysema, chronic obstructive pulmonary disease (COPD), cystic fibrosis)
- ☐ Diabetes
- ☐ Chronic heart disease
- ☐ Chronic liver disease
- ☐ Chronic inflammatory disease (e.g., lupus erythematosus, rheumatoid arthritis, psoriasis, eczema, Crohn's disease, ulcerative colitis)
- ☐ Cancer
- ☐ Immunosuppressing condition (e.g., organ transplantation, HIV-infection, cancer chemotherapy, radiotherapy, congenital immunodeficiency)
- ☐ Chronic renal disease and/or dialysis
- ☐ Asplenia or hyposplenia (absent or defective spleen)
- ☐ Neurologic problem (e.g., Guillain-Barré syndrome, multiple sclerosis)
- ☐ Hematologic problem (e.g., anemia, hemoglobinopathy, bleeding problems)
- ☐ Alzheimer disease or any other dementia
- ☐ Another long-term health condition, specify: ____________________
- ☐ No symptoms and no medicine for these conditions in the past 12 months

### Example of questions S2. Example of the social contact diary

**DATE DAY 1**

| ☐☐☐☐ | ☐☐ | ☐☐ |
|---|---|---|
| Year | Month | Day |

**List of persons with whom you have been in contact during this first day, from 5 am today to 5 am tomorrow morning.**

| Initials or nickname | Age (or age group) of the contacted person | Sex | | Person's relation with yourself | | | | Place of contact (*check all that apply*) | | | | | | Total duration of contact with the person | | | | | Did you touch his/her skin? | | How often do you have contact with this person in general? | | | | | Ethnicity of this person | | | | | |
|---|---|---|---|---|---|---|---|---|---|---|---|---|---|---|---|---|---|---|---|---|---|---|---|---|---|---|---|---|---|---|---|
| | | Female | Male | Household member | Family member not living in household | Friend / Colleague | Other | Home / Car / Private place | Work | Kindergarten / School / College / University | Public transport | Leisure | Other | less than 5 min | 5 - 14 min | 15 - 59 min | 1h - 4h | more than 4 h | Yes | No | Daily or almost daily | A few times a week | A few times a month | A few times a year or less | First time | White | Black | Asian | Hispanic / Latino | Other | Don't know |
| ____ | ☐☐(-☐☐) | ☐ | ☐ | ☐ | ☐ | ☐ | ☐ | ☐ | ☐ | ☐ | ☐ | ☐ | ☐ | ☐ | ☐ | ☐ | ☐ | ☐ | ☐ | ☐ | ☐ | ☐ | ☐ | ☐ | ☐ | ☐ | ☐ | ☐ | ☐ | ☐ | ☐ |
| ____ | ☐☐(-☐☐) | ☐ | ☐ | ☐ | ☐ | ☐ | ☐ | ☐ | ☐ | ☐ | ☐ | ☐ | ☐ | ☐ | ☐ | ☐ | ☐ | ☐ | ☐ | ☐ | ☐ | ☐ | ☐ | ☐ | ☐ | ☐ | ☐ | ☐ | ☐ | ☐ | ☐ |
| ____ | ☐☐(-☐☐) | ☐ | ☐ | ☐ | ☐ | ☐ | ☐ | ☐ | ☐ | ☐ | ☐ | ☐ | ☐ | ☐ | ☐ | ☐ | ☐ | ☐ | ☐ | ☐ | ☐ | ☐ | ☐ | ☐ | ☐ | ☐ | ☐ | ☐ | ☐ | ☐ | ☐ |
| ____ | ☐☐(-☐☐) | ☐ | ☐ | ☐ | ☐ | ☐ | ☐ | ☐ | ☐ | ☐ | ☐ | ☐ | ☐ | ☐ | ☐ | ☐ | ☐ | ☐ | ☐ | ☐ | ☐ | ☐ | ☐ | ☐ | ☐ | ☐ | ☐ | ☐ | ☐ | ☐ | ☐ |
| ____ | ☐☐(-☐☐) | ☐ | ☐ | ☐ | ☐ | ☐ | ☐ | ☐ | ☐ | ☐ | ☐ | ☐ | ☐ | ☐ | ☐ | ☐ | ☐ | ☐ | ☐ | ☐ | ☐ | ☐ | ☐ | ☐ | ☐ | ☐ | ☐ | ☐ | ☐ | ☐ | ☐ |
| ____ | ☐☐(-☐☐) | ☐ | ☐ | ☐ | ☐ | ☐ | ☐ | ☐ | ☐ | ☐ | ☐ | ☐ | ☐ | ☐ | ☐ | ☐ | ☐ | ☐ | ☐ | ☐ | ☐ | ☐ | ☐ | ☐ | ☐ | ☐ | ☐ | ☐ | ☐ | ☐ | ☐ |
